# A transgenerational mutational signature from ionizing radiation exposure

**DOI:** 10.1101/2023.11.20.23298689

**Authors:** Fabian Brand, Hannah Klinkhammer, Alexej Knaus, Manuel Holtgrewe, Leonie Weinhold, Dieter Beule, Kerstin Ludwig, Prachi Kothiyal, George Maxwell, Markus Noethen, Matthias Schmid, Karl Sperling, Peter Krawitz

## Abstract

The existence of transgenerational effects of accidental radiation exposure on the human germline remains controversial. Evidence for transgenerational biomarkers are of particular interest for populations, who have been exposed to higher than average levels of ionizing radiation (IR). This study investigated signatures of parental exposure to IR in offspring of former German radar operators and Chernobyl cleanup workers, focusing on clustered de novo mutations (cDNMs), defined as multiple de novo mutations (DNMs) within 20 bp. We recruited 110 offspring of former German radar operators, who were likely to have been exposed to IR (Radar cohort, exposure = 0-353 mGy), and reanalyzed sequencing data of 130 offspring of Chernobyl cleanup workers (CRU, exposure = 0-4,080 mGy) from Yeager, et al. In addition, we analyzed whole genome trio data of 1,275 offspring from unexposed families (Inova cohort). We observed on average 2.65 cDNMs (0.61 adjusted for the positive predictive value (PPV)) per offspring in the CRU cohort, 1.48 (0.34 PPV) in the Radar cohort and 0.88 (0.20 PPV) in the Inova cohort. This represented a significant increase (*p* < 0.005) of cDNMs counts, that scaled with paternal exposure to IR (*p* < 0.001). Our findings corroborate that cDNMs represent a transgenerational biomarker of paternal IR exposure.

## Introduction

Transgenerational effects of ionizing radiation (IR) in the offspring of former radar operators are a stated concern of several European armed services ^1^. Investigation of such effects is warranted in order to design effective preventive measures, and optimize health monitoring of military personnel and their offspring.

The potential of transmission of radiation-induced genetic alterations to the next generation is of particular concern for parents who may have been exposed to higher doses of IR and potentially for longer periods of time than considered safe. To address the concerns articulated by former radar soldiers, the German Ministry of Defense initiated this study with the goal of investigating the transgenerational effects of IR in offspring of exposed parents. Some tasks, primarily mechanical calibration work on radar units during live operations, have been recognized as an occupational health hazard by the German government ^1^.

Previous studies into the transgenerational effects of IR on human DNA have investigated populations affected by nuclear weaponry, and the Chernobyl nuclear reactor incident of 1986 ^1–8^. Recent studies in a small cohort of British nuclear test veterans found no enrichment in chromosomal aberrations, de novo mutations, or structural variation in the offspring of exposed soldiers ^2,3^. However, a transgenerational effect could not be excluded, due the limited cohort size and the low and uncertain estimates of IR exposure ^2,3^. Two studies of the impact of the Chernobyl incident found a twofold increase in mutation rates at minisatellites, microsatellites, and tandem repeat loci in the offspring of former clean-up workers ^5,9^. However, the small cohort sizes, potential confounders, and the limited number of loci that were examined have led to ongoing debates concerning the statistical significance of these findings ^10–12^. Yeager, et al. recruited a cohort of 105 individuals who had been exposed to IR following the 1986 Chernobyl nuclear accident and their 130 offspring (born 1987-2002). The parents had either been inhabitants of the town of Pripyat at the time of the accident, or had been employed as liquidators responsible for guarding or cleaning up the accident site ^8^. No elevated mutation rates for isolated DNMs were detected ^8,13^. Furthermore, the authors found no enrichment of C>T mutations within 47 kb intervals, which has been hypothesized as an indicator of hyper-mutability of single-strand intermediates during the repair of double-strand breaks, but no smaller mutation clusters were analyzed ^14,15^. Therefore, to date, no definite transgenerational mutational signature of IR exposure has yet been identified.

Identification of a potential transgenerational mutational signature for IR exposure requires a detailed understanding of the impact of IR on human DNA in the germline. When IR interacts with DNA the energy transfer may directly cause. a variety of DNA lesions, such as strand breaks, oxidized bases, loss of bases ^16–18^. However, the primary pathway for IR-induced DNA damage is often indirect; IR generates reactive oxygen species (ROS) through the ionization of nearby water molecules in the cell ^16–18^. These ROS then induce a variety of DNA lesions, including oxidized bases, base losses, double strand breaks (DSBs), single strand breaks (SSBs), with DSBs being the most detrimental to DNA structure. The repair of DSBs involves two main mechanisms: 1) Homologous Recombination Repair (HRR), a process that involves a homologous template; and 2) Non-Homologous End Joining (NHEJ), a process in which broken DNA ends are ligated without a template ^19^. In germline cells, particularly during spermatogenesis, HRR plays a critical role in maintaining genomic integrity, whereas NHEJ, despite being more common, is more likely to introduce errors ^20^. Due to its error-prone nature, NHEJ in germline cells can result in complex, ROS-induced lesions, which are turned into mutations, within short genomic regions ^17,21^. Consequently, these lesions may contribute to genomic instability and cause cell death or persist through cell division and, particularly in germline cells, be passed on to future generations, representing a potential signature of IR parental exposure ^21,22^. Importantly, DNA repair is less efficient in spermatids and mature spermatozoa which, consequently, show the highest radiosensitivity of all stages of spermatogenesis ^23,24^.

Recent research in mice has provided compelling evidence that clustered de novo mutations (cDNMs) within short DNA segments (<20 bp) can increase following paternal exposure to IR, with the magnitude of this effect being dose-dependent, particularly in hematopoietic stem cells ^25–27^. To investigate this finding in humans, in 2018, our group performed a small WGS pilot study of 18 offspring of former radar operators from the German military ^7^. The analyses identified an increased mean number of cDNMs (then called multi-site de novo mutations (MSDNs)), and cases with exceptionally high cDNM rates. The analyses also identified two translocations, which had resulted from neighboring mutations. The results of this pilot study suggest that cDNMs may represent a signature of IR-induced DNA damage in humans.

Accurate identification of DNMs and cDNMs in current generation WGS data has to account for natural and technical biases. Parental age at conception is a significant and known confounder for the number of *de novo* mutations in their offspring ^14,28,29^. Paternal age is associated with an increase of isolated DNMs, averaging at roughly 1-2 DNMs per year of paternal age at conception of the child. In addition, the maternal age has primarily been implicated with rarer, more clustered mutations, showing a significant enrichment in 10k bp wide clusters ^14,30^. WGS data enables the analysis of DNMs and cDNMs all over the human genome, but due to lower specificity in DNM calling many regions with repeats or regions with low genomic complexity have been excluded from earlier studies ^7,25,28^. It is particularly hard to distinguish DNMs from sequencing errors in these regions, and the difficulty to capture these regions using PCR primers makes validation of potential DNM and cDNM calls challenging. The DNA source material and sequencing device that generates the data also have an influence on the quality and accuracy of DNM calls ^31^.

The aim of the present study was to determine whether the signature of IR-induced clustered DNA lesions was detectable in the offspring of fathers with a history of probable IR exposure. The analyses were conducted using a newly recruited cohort of former German-military radar operators, their wives, and offspring (Radar cohort), as well as WGS data accessed from Yeager et al. 2021 (CRU cohort), and from a previously reported cohort of individuals with no history of exposure to IR (Inova cohort). Herein, we describe how we tested whether cDNMs are a transgenerational biomarker of prolonged paternal exposure to IR. We sequenced data for the Radar cohort and performed a new joint variant calling analysis together with the CRU and Inova data in order to confirm the known paternal age effect in all three cohorts. Afterwards, we used negative binomial regression models to ascertain differences in the number of cDNMs per sample in each cohort and to associate them with the likely exposure of their fathers.

## Materials and Methods

### The Radar cohort

The study at hand was commissioned by the *“Bundesamt für Ausrüstung, Informationstechnik und Nutzung der Bundeswehr”* (Federal Office of Bundeswehr Equipment, Information Technology and In-Service Support, BAAINBw) to further improve the compensation for former radar personnel of both German armies (BT Drs. 18/9032). Recruitment for the present Radar cohort was conducted between 2019 and 2021. Former radar soldiers were approached by the study team via advertisements in relevant magazines and online forums, and through the “Bund zur Unterstützung Radargeschädigter e.V.”, a support group for potentially IR-exposed former German radar operators. The primary inclusion criterion for the present study was a history of exposure to high dose IR when servicing radar installations during live operations, as judged by an independent expert on the basis of a self-report questionnaire sent to each potential participant. In total, 80 former radar operators from the West or East German armies (Bundeswehr and Nationale Volksarmee, NVA) were included in the present study, together with their wives (n = 80) and offspring (n = 110) (Supplemental Table S1) ^1,7^. Even though the German government spent significant efforts in investigating health effects and occupational risks following long time service with radar units, and despite the fact that soldiers serving at unprotected radar installations have a higher risk to develop certain cancers, reliable data on the damage caused by the stray radiation from these devices is very limited ^1,6^.

### Ethics Statement

Ethical approval for the present study was obtained from the ethics committee of the Medical Faculty of the University of Bonn (Ethikkommission der Medizinischen Fakultät Bonn). All participants from the Radar cohort, i.e. the former soldiers, their wives, and their offspring, provided written informed consent for the present analyses prior to inclusion. All participants of the CRU and Inova cohorts had provided written informed consent for the use of their data by other research groups within the context of the respective original investigation. The present analyses were performed within the guidelines specified in the informed consent documentation for all three cohorts, and within the limits of the approval granted by the ethics committee of the Medical Faculty of Bonn. All study procedures were conducted in accordance with the principles of the Declaration of Helsinki.

### Estimation of IR dose in the Radar cohort

Retrospective estimations of IR dose for the Radar cohort were made at the Radiation Measurement Facility (Strahlenmessstelle) of the German Federal Armed Forces (Bundeswehr) ^32^. For this purpose, the military service record of each Radar participant was accessed. In estimating IR dose, two factors were considered. The first factor was the role and duties of the given Radar cohort participant, and the period of time for which they had served in a radar unit of the West or East German army (Supplemental Material 1.2). The second factor was the radar device that had been in active service at the time of the participant’s military service. Dose estimations were based on: 1) historical measurements, which had been taken from common radar devices by the military at the time the device had been in active service; or 2) measurements of the emissions of out-of-service radar devices that were reconstructed for the purpose of retrospective assessment. For each radar device, potential sources of stray radiation were determined in order to establish a realistic base rate of IR emission during service. Even though the retrospective dose assessment was carried out with great care, it is likely that there are errors introduced by both aforementioned factors that formed the basis for the dose estimation. Section 1.2 of the Supplemental Materials gives a detailed account of the procedures for retrospective dose estimation.

### Whole Genome Sequencing Data

For all participants of the Radar cohort, sequencing was performed at the NGS core facility of the West German Genome Center (WGGC) in Bonn to a minimum whole genome coverage of 30X. WGS was performed according to the standard protocols on an Illumina NovaSeq device (Supplemental Material 1.3). To ensure that data generated on HiSeq devices (i.e. Inova and a subset of CRU) and sequences that were generated by the newer generation NovaSeq devices (i.e. Radar and the remainder of CRU) were comparable, and that the different read lengths did not induce confounding errors, three families from the Radar cohort were sequenced on both devices.

WGS data from the cohort of Yeager, et al. were accessed under dbGAP accession number phs001163.v1.p1, and all parent-offspring trios were downloaded from dbGAP. All offspring in the CRU cohort were older than 18 years and apparently healthy, only 13 of them were conceived at the time of the reactor accident, most of them many years later ^33^.

In addition to the two case cohorts described above, we accessed trio WGS control data from the Inova cohort of Wong et al. ^28^. The Inova cohort comprises 1,214 familial trios (Inova, Supplemental Material 1.1.2, Supplemental Table S1), with no recorded history of exposure to non-naturally occurring IR ^28,30,34^. This WGS data was also used to analyze the effect of parental age on isolated DNMs in the germline and genetic effects on preterm births ^28,30,34^. We downloaded the Inova WGS data from an AWS S3 bucket.

### Variant Calling and Quality Control

For the Radar and Inova cohorts, variant calling was performed using Illumina DRAGEN v3.6.3 in the Amazon web services cloud (Region: Ireland). For the CRU cohort, data were processed on an on-premises computing cluster using NVIDIA Parabricks (Supplemental Material 1.4.1). For variant analysis, all data were aligned to the GRCh37 reference genome, and joint variant calling was performed using GLnexus v1.3.1 on a total of 4,337 whole genome sequences ^35^. For all samples and cohorts, we subsequently performed exhaustive quality control checks (Supplemental Material 1.4.3, Supplementary Figure S5, S6). These included sex and ancestry controls using Peddy, as well as assessments of contamination, sequencing depth and variants (e.g. transition-transversion ratio), in order to control for any technical bias that may have arisen secondary to differences in sequencing technology, chemistry, or other forms of error ^36–40^. Further detailed information about the variant calling efforts is present in the Supplemental Material, Section 1.4.1.

#### Detection of de novo and clustered de novo mutations in all parent offspring trios

To detect DNMs first and later cDNMs, a set of filters was applied to all three cohorts in parallel. For each of the three cohorts, the output of the variant calling pipeline formed the basis for the detection of DNMs and cDNMs. For each parent-offspring trio Python3 and Hail v0.2.89 were used to find potential DNM candidates and to refine this call set to clusters, as based on a window size of 20 bp ^41^. The detection of DNMs was based on heuristics, including the score calculated by “hl.de_novo” (> 0.85); sequencing depth at the site (>10); parental genotypes and read data (< 2 reads featuring the variant allele); the allele count in all samples (AC <= 1); and other criteria (Supplemental Material 1.4.2). Supp. Table S2 shows the filtering settings that were used to detect DNMs. The most stringent filter employed in the detection of DNMs was the AC = 1 filter, whereby any variant with an *AC* > 1 in the combined cohort (Radar, CRU, Inova) was discarded. This filter removed all familial variants as well as DNMs and cDNMs with a high population allele frequency whose origin was thus unlikely to have been radiation^42^. To ascertain the quality of all *de novo* calls made in the Radar cohort, replication of each DNM call was attempted using Graphtyper. A concordance rate for DNM calls in each sample was then calculated ^43–46^.

#### Phasing

To determine the parental gamete of origin, read-based phasing of DNMs with informative variants was used (Supplemental Material, Section 1.4.4) ^47^. Since clustered DNMs can extend over several base pairs, not all lesions in a cluster can necessarily be phased. If read-based phasing suggested the paternal or maternal germline based on the information from at least one lesion we assumed this origin for the whole cluster. Clusters where multiple DNMs showed differing evidence for paternal or maternal origin were called contradictory cDNMs (Supplemental Material, Section 1.4.4). In addition to read-based phasing, for a subset of cDNMs in the Radar cohort, the parental origin was also determined using Sanger and PacBio long-read sequencing. We resequenced a phase informative single nucleotide polymorphism (SNP) alongside each cluster, which uniquely identified the parental origin of each cluster.

#### cDNM Window Size

cDNMs are defined as genomic regions where at least two *de novo* mutations occur within 20 bp distance of each other. A window size of 20 bp was selected as the cutoff for cDNMs, since we hypothesized that the ROS-induced DSBs are the primary driver of radiation induced cDNMs in the human germline. ROS affects human DNA in a range of only 4-6nm, and 20 bp is the interval size used for these clusters in previous investigations^4^. Clusters of this size have also earlier been implicated in gonadal exposure to IR in humans and mice ^7,8,18^. We also assessed different cluster window sizes that have been used in the literature in a sensitivity analysis (10 bp, 30 bp, 10k bp, 47k bp) by recalling all cDNMs with the given window size ^2,8,14,48^. Cluster sizes in the range of 10k bp to 100k bp have previously been connected with the maternal age effect by some studies, but no association with ionizing radiation has been shown thus far ^8,14,30^.

#### cDNM Calling

Using the selected DNMs, which passed all filtering criteria, *de novo* mutation clusters were assembled using a trivial algorithm, whereby DNMs were added to a single cluster, if the distance to their direct predecessor on the same chromosome was < 20 bp. A cluster with > 2 lesions can span a total size larger than 20 bp.

### Validation of cDNMs in the Radar Cohort

Since cDNMs have a higher false positive rate than germline mutations and isolated de novo events, all cDNMs in the Radar cohort were validated by at least one of three methods using a three-step iterative approach (Supplemental Material 1.5, Supplemental Figure S7). This involved the use of Sanger and PacBio sequencing data to derive criteria for identifying true and false positive clusters, as based on the IGV Browser visualization. For both Sanger and PacBio sequencing, primers were first designed using Primer3, followed by manual optimization based on available short-read sequence data ^49,50^. However, due to the complex nature of the genomic regions in which the candidate cDNMs were located, many were not validated in subsequent experiments.

Sanger sequencing was conducted on a subset of 71 potential cDNMs. These spanned the following categories: tandems (n=14); GG>TT tandems (n=15); indels (n=21); large cDNMs (involving more than three lesions; n=5); and cDNMs located within repetitive regions (n=16). Of the 71 analyzed clusters, interpretable results were obtained for 44. Specifically, 5 clusters were true positives, 39 clusters were false positives, and the status of 27 clusters remained undetermined due to sequencing challenges. Notably, none of the GG>TT tandem clusters achieved validated true positive status, emphasizing the difficulty in accurately sequencing certain mutation types in these regions. From this dataset, a set of guidelines were established for ascertaining which potential cDNMs were true or false positive calls (Supplemental Material 1.5, Supplementary Figure S7). The data from the final validation callset was used to establish the positive predictive value (PPV) of cDNM calls on the Radar cohort. In the statistical analysis, the PPV was used exclusively for a downsampling simulation (Supplemental Material 1.6.6).

### Statistical Analysis

To ascertain potential statistical differences in the count data in this study (e.g. number of DNMs or cDNMs per offspring), generalized linear models were used (Supplemental Material 1.6.2). Under Bonferroni correction, the significance level was set at 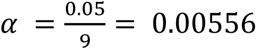 for nominal p-values *p*_*nom*_, or p-values were adjusted to *p*_*adj*_ = 9*p*_*nom*_.

Since parental age at conception is a known and significant confounder for all analyses including DNMs, correction for the paternal and maternal age was performed in all analyses, unless otherwise stated. Since paternal and maternal age are highly correlated (Pearson-R: 0.71, *p* < 5 ⋅ 10^−100^), the age of the father at conception was used as a proxy for the effect of parental age, which includes both maternal and paternal age effects (Supplemental Material 1.1.4, Supplemental Figure S2). For models that did not incorporate paternal age directly, age matching was used to control for this confounder (Supplemental Material 1.6.1), by selecting subcohorts of Radar, Inova and CRU with homogenous age distributions. The age matching procedure downsampled Inova and CRU to the same or n-times (*n* ≥ 1) the size of the smallest cohort (Radar, n = 110). This was achieved by computing the minimum-weight bipartite matching between node sets representing the individual samples of any two cohorts. Two nodes in the graph, representing offspring in either cohort, were connected by an edge, whose weights were set to the sum of the age differences of the mothers and fathers. The minimum weight bipartite matching in this graph is then the subcohort of Inova and CRU respectively, that minimizes the age difference indicated by the edge weights in the graph (Supplemental Material 1.6.2).

## Results

### Estimated IR dose

Because some soldiers served in military roles that probably did not result in elevated levels of exposure, and due to the challenging retrospective dose estimations, the dose estimations remained inconclusive for the majority of soldiers (*n*_*exposed*_ = 22, *n*_*no* *exposure*_ = 55, *n*_*no* *documents*_ = 3, Supplementary Figure S3, Supplementary Table S10, Supplementary Material “Bericht S209/20”). We call the offspring of soldiers with an estimated dose of > 0 mGy the Exposed subcohort, while the Unexposed subcohort was comprised of all children of fathers that were deemed unlikely to be exposed.

Dose estimations for the Radar cohort were performed after the recruitment phase ended and the average estimated IR dose in the total Radar cohort was 9.21 (± 53.33) mGy (median = 0 mGy). In the subgroup of radar technicians with a dose estimation of > 0 mGy (n = 22), the average was *μ* = 34.35 (±99.77) mGy (*median* = 0.0021 mGy) (Figure 1a) ^32^.

**Figure 1:**
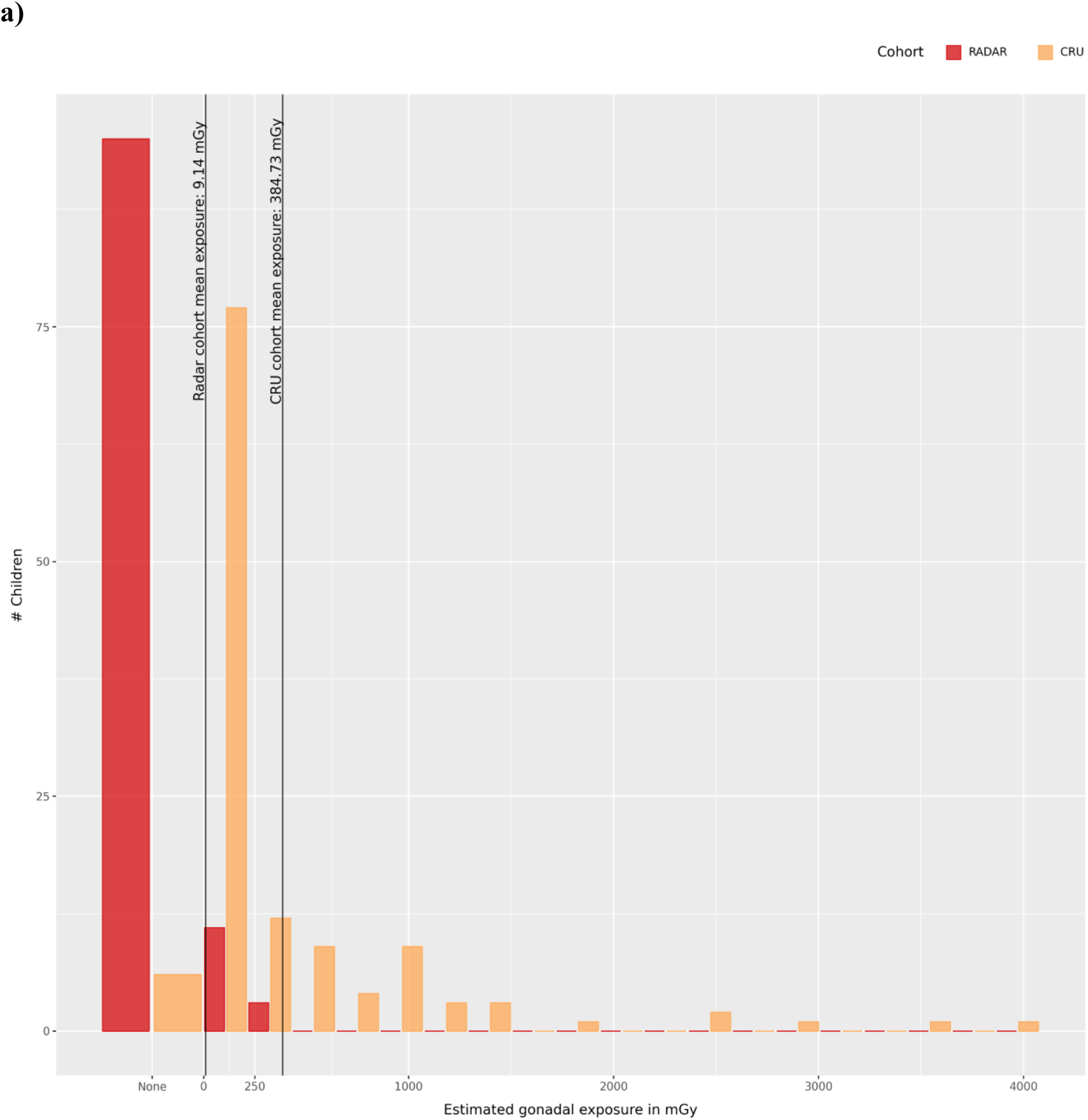
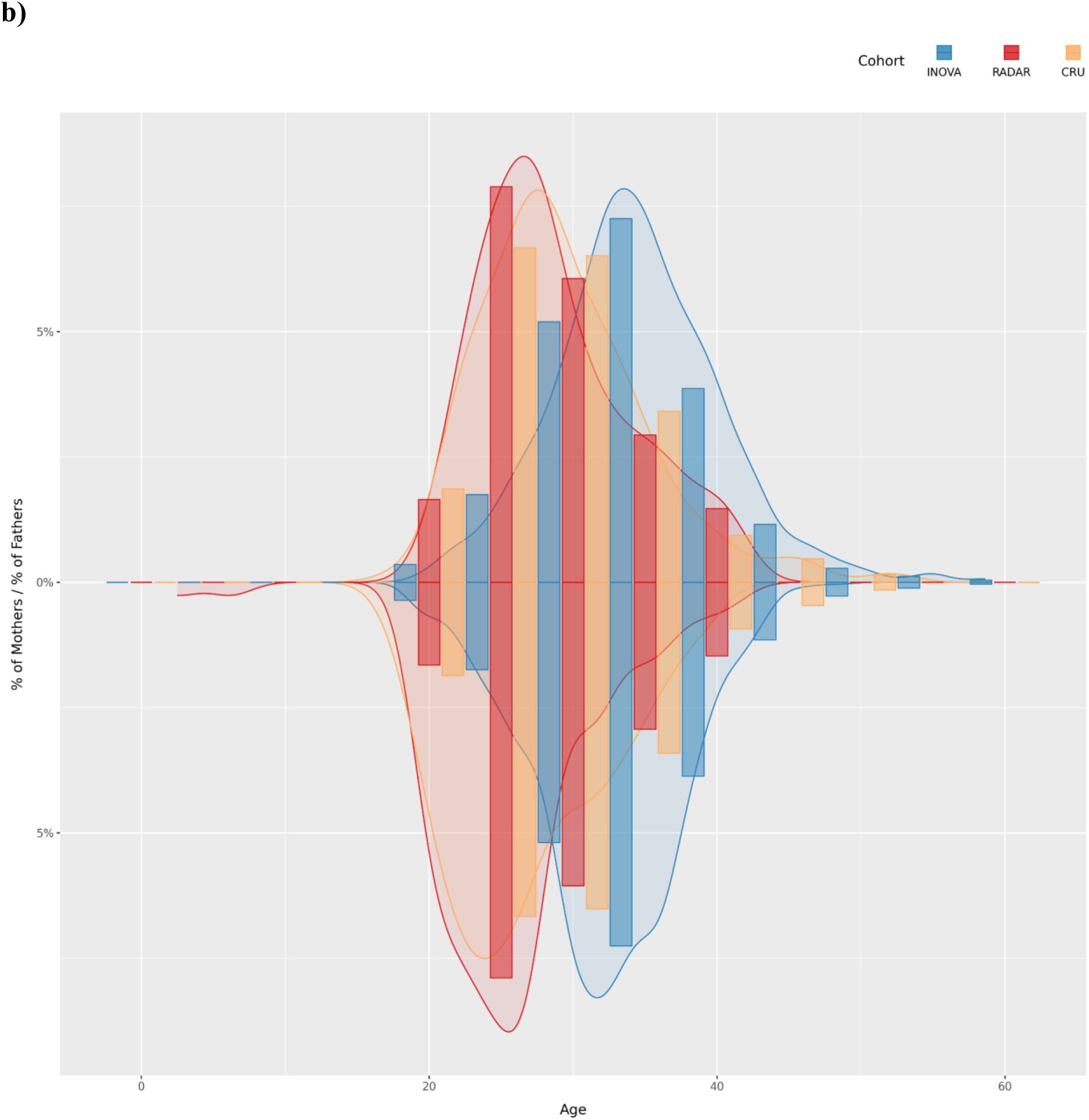
Study Cohorts. **a)** Distribution of paternal exposure for the Radar and CRU cohorts. The maximum exposure observed in the Radar cohort is 353 mGy, and 4,079 mGy in the CRU cohort. In the Radar cohort, 77 out of 110 children were born to soldiers with a dose estimation of 0 mGy, 30 to soldiers with a valid exposure estimation > 0 mGy and for the father of 3 offspring, the dose estimation could not be made. **b)** Age distribution of the three study cohorts. Due to the large differences in cohort size, the y-Axis indicates the percentage of the total cohort size. Values in the bottom half of the y-Axis show the distribution of maternal age, and values in the top half show the distribution of paternal age. On average, fathers in the Inova cohort were >5 years older compared to fathers in the Radar and CRU cohorts, and mothers were >5.5 years older on average.

For the CRU cohort, dose estimations were accessed from the data published by Yeager, et al.^8^. On average, fathers in the CRU cohort were exposed to *μ* = 365.42 (±684.55) mGy (*median* = 29) of IR (Figure 1a, Supplementary Material, Section 1.1.3, Supplementary Figure S1).

### Analysis of de novo mutations

After obtaining all necessary data, we processed all samples using the equivalent bioinformatics pipelines, as detailed in the Materials and Methods section. Before computing the set of DNMs, we confirmed that all quality control checks passed, in particular that there was no substantial difference in the whole genome coverage between the cohorts, and that the pedigree in all cohorts matches the reported family structure (Figure 1b, Supplementary Figure S4). In accordance with the literature, the number of isolated DNMs increased by 2% per year of paternal age in all three cohorts (Figure 2, Supplementary Table S5), which translates to an accumulation of 1-2 mutations per year of age of the father ^28,29,51,52^. In the age-matched analyses, the rate of isolated DNMs per generation was: (i) Inova, 72.67 (18.15, median = 79); (ii) CRU, 65.43 (13.57, median = 65); (iii) Radar, 67.95 (17.25, median=64) (Supplemental Tables S1, S3). For Inova and CRU, these rates are comparable to the values reported in the original studies. No significant difference in the rate of isolated DNMs per generation was found between the three cohorts (Supplemental Material, Section 1.6.3, Supplementary Table S4) ^8,28^. None of the datasets showed a bias towards specific nucleotide exchanges, as has been reported previously for certain generations of sequencing devices (Supplemental Figure S8) ^31^. Our bioinformatic replication using the Graphtyper algorithm yielded a concordance of ≥ 88 % for DNMs in the radar cohort. The sequencing replicates of three families yielded a PPV of 90.2% for the DNM calls made on the NovaSeq, assuming calls from the HiSeq as ground truth. (Supplementary Table S14).

**Figure 2:**
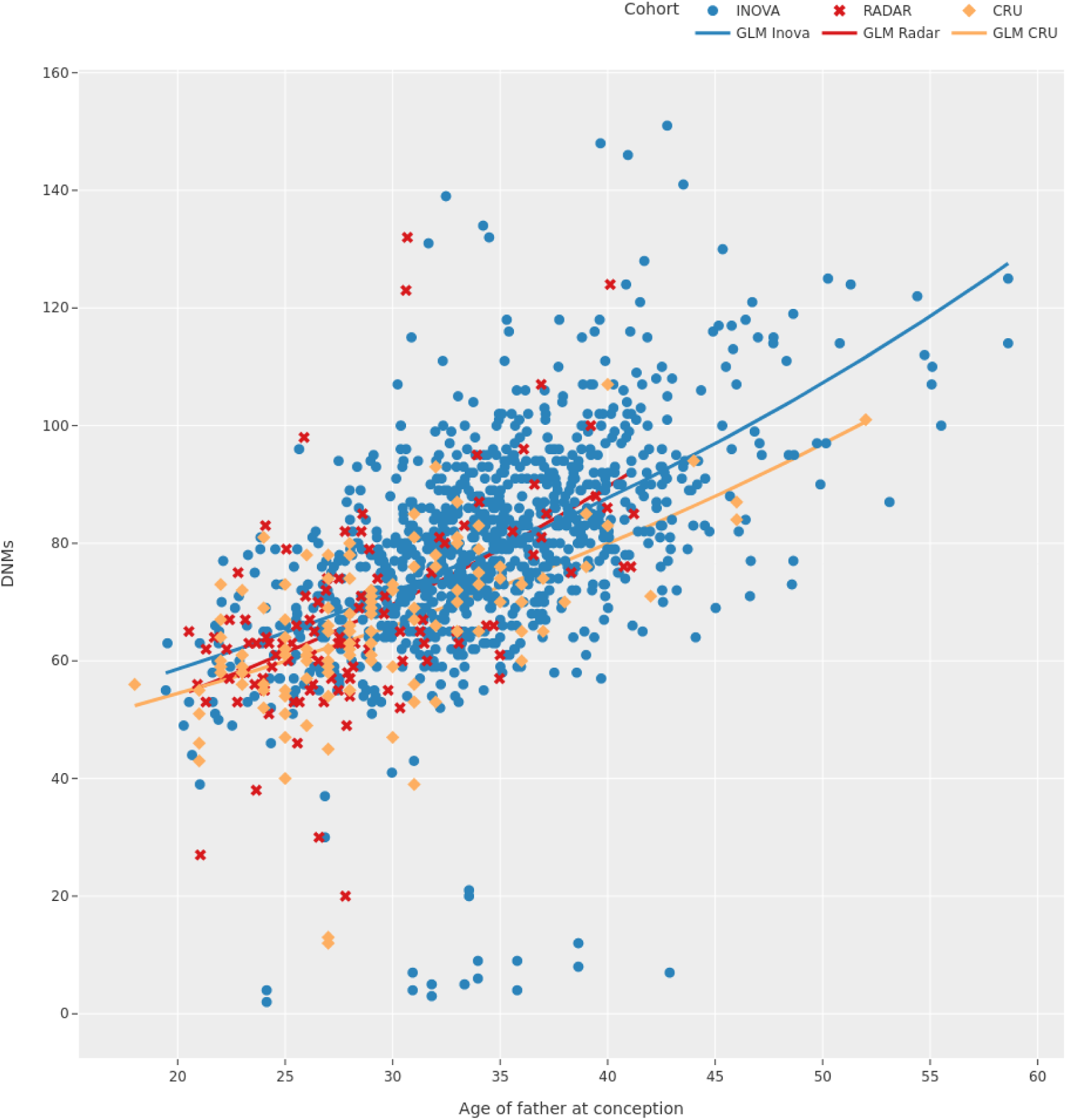
Paternal Age Effect. Paternal age effects computed by a negative binomial regression model to estimate the number of DNMs according to the paternal age at conception for the offspring in each cohort. When fitting this model, no age matching was applied to the data. Therefore, on average, the parents are older in the Inova cohort. Nevertheless, the paternal age effect for each of the cohorts is approximately 2%, which results in an increase of ∼1 DNM per year of paternal age.

### Analysis of clustered de novo mutations

We continued our analysis by filtering for loci with multiple lesions (clustered DNMs, cDNMs). In total, 1,989 cDNMs were detected in 1,515 offspring (*n*_*Inova*_ = 1275, *n*_*Radar*_ = 110, *n*_*CRU*_ = 130). These mutations were enriched in offspring of irradiated fathers in the Radar cohort (*μ* = 1.48 ± 1.72, *median* = 1) and in the CRU cohort (*μ* = 2.65 ± 2.65, *median* = 2) compared to the Inova cohort (*μ* = 0.88 ± 0.98, *median* = 1) (Supplemental Table S6). In offspring from the CRU cohort, the median number of clustered DNMs was two, which was twice as many as that detected in the age-matched subset of the Inova cohort. A negative binomial regression model confirmed that the estimated number of cDNMs in the Inova cohort was less than in the Radar or CRU cohort (*n* = 110, 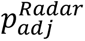 = 0.045, 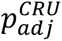 < 1 ⋅ 10^−3^; Figure 3, Supplemental Table S7). These differences were more prominent when the Radar cohort was divided into the Exposed and Unexposed subcohorts, where the children of exposed parents showed a higher number (Exposed = 1.72, Unexposed = 1.39) of cDNMs on average (Supplemental Material, Section 1.6.8, Supplementary Figure S10).

**Figure 3:**
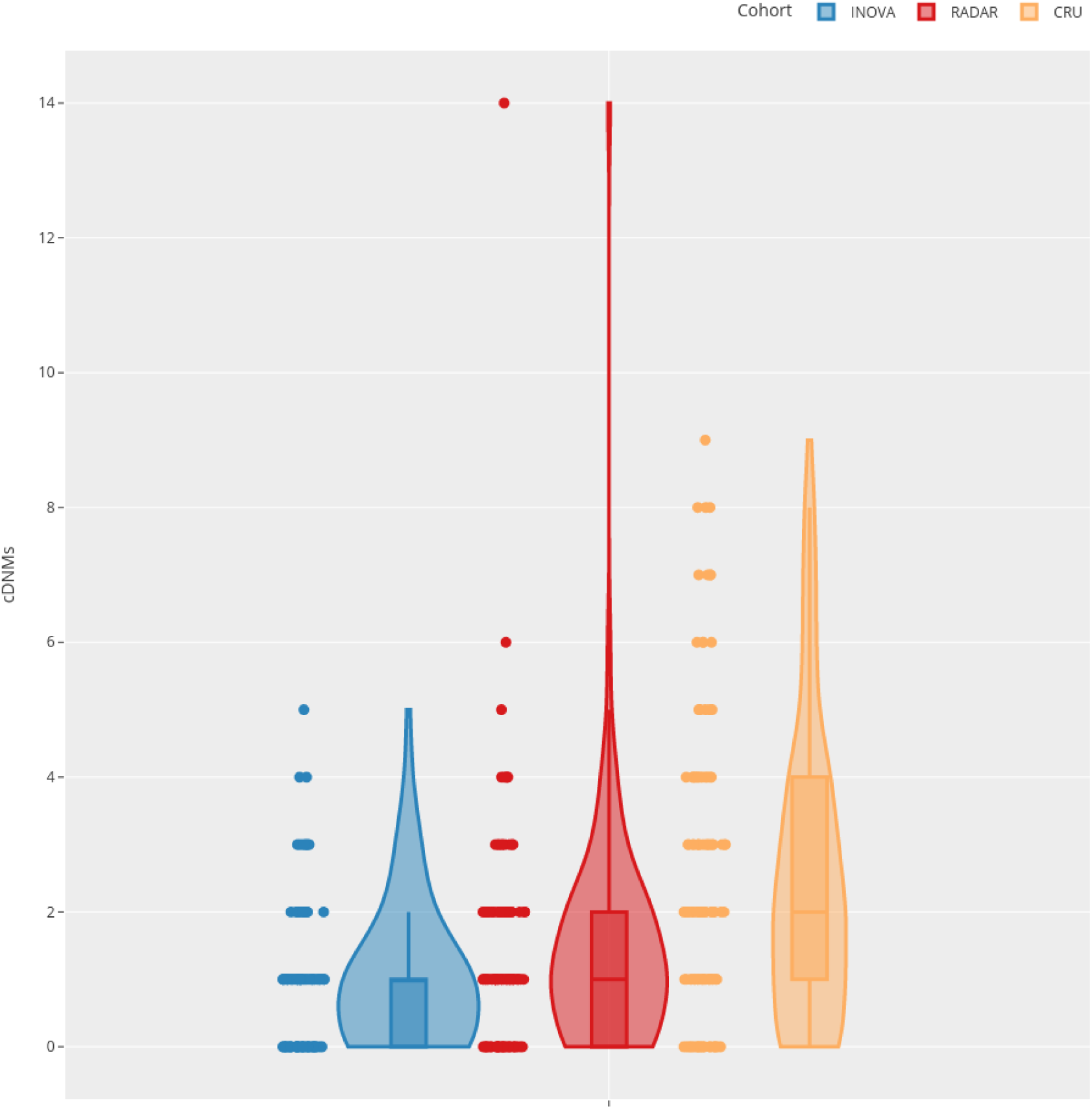
Number of cDNMs per sample. Violin plot of the number of clustered de novo mutations (cDNMs) per sample, as grouped according to cohort. The width of the violin at each integer value of the y-axis indicates the number of samples and their respective number of cDNM clusters, without correcting for the PPV of 0.23. Our simulation experiments controlling for the effects of this PPV on the statistical tests are presented in Supplemental Table S8. The box plot for each cohort is included inside the respective violin to display the quartile ranges and median number of cDNMs per sample in the respective cohort. On average, the age matched analysis detected 0.88 (±0.98; *median* = 1; *ppv* − *adjusted* = 0.20) cDNMs in the Inova cohort, 1.48 (±1.72; *median* = 1; *ppv* − *adjusted* = 0.34) in the Radar cohort, and 2.65 (±2.19; *median* = 2; *ppv* − *adjusted* = 0.61) in the CRU cohort.

We also found cDNMs to be significantly increased in offspring of irradiated fathers for 10 bp and 30 bp windows in our sensitivity analysis. In contrast, the larger window sizes led to a reduction in the difference in the number of cDNMs between the cohorts and a substantial drop in *p*_*nom*_(Supplemental Material, Section 1.6.12, Supplemental Figures S13, S14).

### cDNM Validation in the Radar cohort

In general, the false positive rate is higher for clustered DNMs ^48^. Our visual inspection criteria were applied to 163 cDNMs in the Radar cohort. Of these, 37 were found to be true positives, 17 of which were also confirmed by PacBio and/or Sanger sequencing data. Therefore, the final PPV for cDNM detection in the Radar cohort was 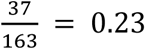 (95% Clopper-Pearson confidence interval 0.17 - 0.30). Notably, in some individuals, none of the detected cDNMs could be validated, including the outlier with 14 cDNMs shown in Figure 3. However, this had no substantial effect on the negative binomial regression model, since this models the median count of cDNMs per offspring, which is robust against outliers. Additionally, simulations accounting for this PPV did not affect the significance of the test results (Supplemental Material, Section 1.6.6, Supplementary Table S8).

All true positive cDNMs identified in the Radar cohort were analyzed with respect to their likely relevance to disease states or their impact on the coding region in general. None was found to have any implications in terms of genetic conditions reported by the study participants (Supplemental Material, Section 1.5.1, Supplementary Data “Clinical Data”).

### Phasing of DNMs and cDNMs

For technical and stochastic reasons, the proportion of DNMs that could be phased varied between the three cohorts. The influencing factors were the distance between DNM and the phase-informative SNP, the coverage in the respective region, the length of the sequencing reads (100bp in the control cohort vs 150bp in both case cohorts), and the distribution of fragment sizes. No inter-cohort differences were observed in the number of isolated DNMs that were attributable to the paternal or maternal alleles (Chi-Squared-Test, Supplemental Material, Section 1.6.7, Supplementary Figure S9, Supplementary Table S9). We did not observe any contradictory cDNM clusters. The parental origin of 26 clusters in the Radar cohort was validated, with 17 clusters of paternal and nine of maternal origin being present. Due to the shorter read length of 100bp in the Inova cohort, no reliable estimate for this ratio in the population could be computed.

### Analysis of radiation exposure

In addition to an increase in the number of cDNMs per sample in the Radar and CRU cohorts, a positive correlation was found between the estimated dose and the number of cDNMs per sample. Using a negative binomial regression model, a significant (*p*_*adj*_ < 0.009) increase in cDNMs was observed per mGy of paternal radiation exposure. The regression model estimated the increase of cDNMs as *f*(*n*) = 1.55 ⋅ *e*^0.0005*n*^ mutations per n mGy, when combining the Radar and CRU cohorts and *β*_*CRU*_ = 0.0005 and *β*_*Radar*_ = 0.0007 when analyzing each cohort separately (Figure 4, Supplemental Material 1.6.10, Supplementary Table S12, Supplementary Figure S12). However, it was not possible to assert statistical significance for this model in either the Radar cohort alone, or for the inverse model, by inferring the paternal IR dose from the cDNM count of the respective offspring (Supplementary Section 1.6.11, Supplementary Table S13). The highest number of DNMs per cluster observed in the three cohorts was eight (Inova), nine (Radar), and 11 (CRU) mutations respectively (Supplemental Figure S11, Supplementary Table S11). An analysis of the distribution of cluster sizes across the three cohorts yielded no statistically significant shift (Supplemental Material, Section 1.6.9).

**Figure 4:**
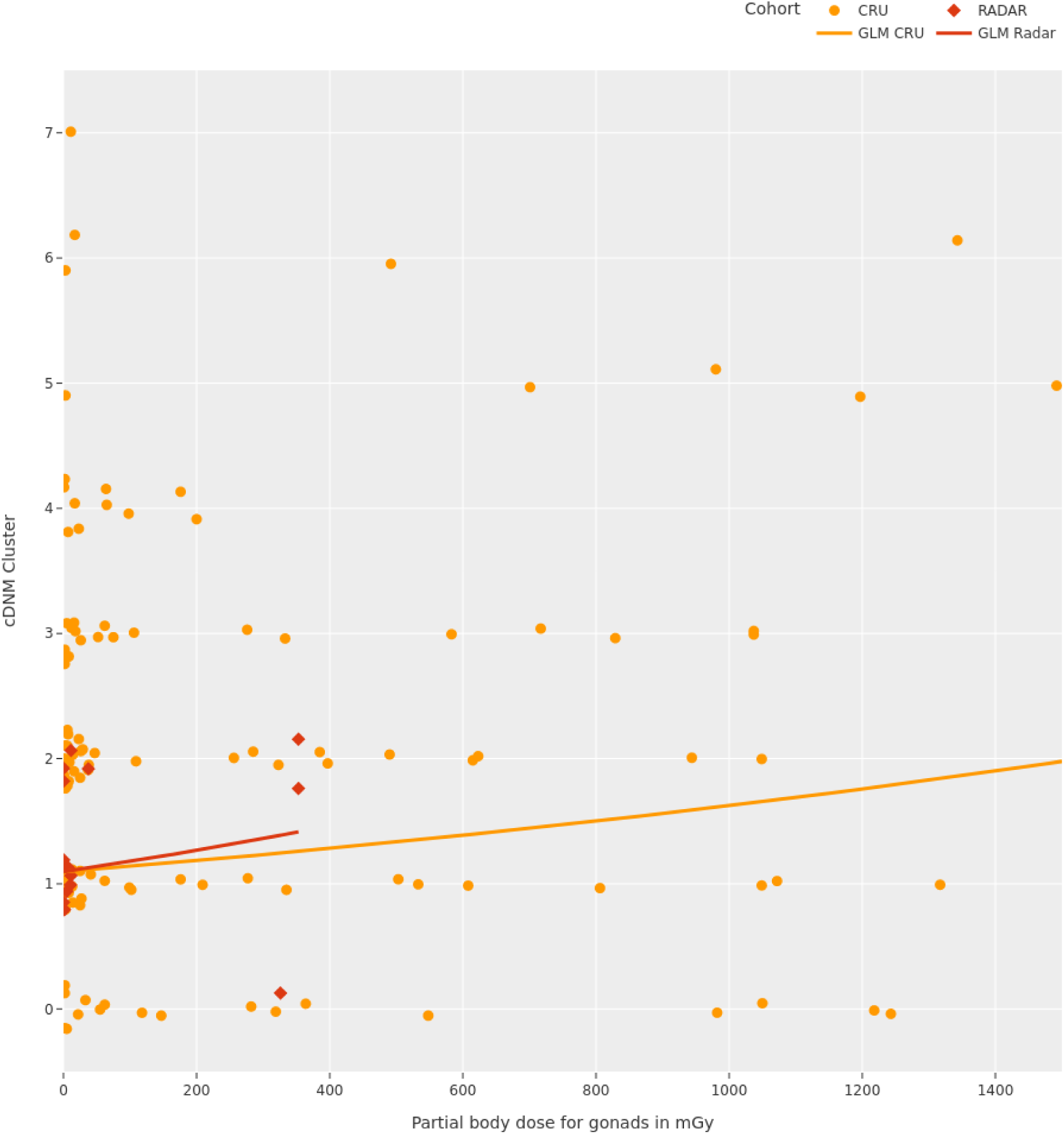
Number of cDNMs per mGy of paternal exposure. Estimation of the number of cDNMs according to the paternal exposure in mGy as computed by a negative binomial regression model with logarithmic link function. Since the accuracy of the negative binomial regression model deteriorates rapidly with larger exposure estimates, the x-axis has been cut off at 1.5k mGy, which means that five samples from the CRU cohort, for which the estimation by the model is very inaccurate, are hidden. The fit shown in this image is conditional on the number of mGy of paternal exposure and the cohort. A model integrating both cohorts into one exposed supercohort is shown in Supplemental Figure S12. This was restricted to the samples from the Exposed subgroup of the Radar cohort, i.e. all offspring of fathers with an estimated exposure of >0 mGy. This restriction in the cohort leads to the exclusion of some offspring with many cDNMs, e.g. the outlier in the Radar cohort in Figure 3.

## Discussion

The question of whether IR confers transgenerational health effects on the human genome has been a topic of research for over 70 years, i.e. since epidemiological studies first investigated the offspring of atomic bomb survivors ^53^. However, the disadvantage of epidemiological analyses is that they require a readout on the phenotypic-level such as malformations ^53^. More recent studies indicate that larger quantities of environmental radiation lead to a higher incidence of cancer and birth defects, emphasizing that there are subtle effects of ionizing radiation on the human germline that have not been captured by earlier studies ^54^. If an analysis is based on phenotypic data alone, an increased rate of *de novo* mutations or other subtle changes may remain unobserved, since most *de novo* mutations are rare and occur in the non-coding part of the DNA. Therefore, the unique capabilities of whole genome sequencing can yield much deeper insights into the consequences of prolonged ionizing radiation exposure on the human genome than possible with earlier sequencing technologies. Yeager, et al. already studied isolated *de novo* mutations and clusters on the scale of kb, which are associated with the repair of double-strand breaks, and did not find a significant increase in mutation rates ^8,28^. Lawrence, et al. and Moorhouse, et al. studied DNMs, structural variants and chromosomal aberrations in offspring of British nuclear test veterans with unclear total radiation exposure. Similar to Yeager, et al., they did not find increased mutation rates in any of their target mutation types, which included DNM clusters with 10 bp or 100 bp size ^2,3^. In mice and somatic cells however, clusters on the single-bp scale were previously implied as a consequence of the secondary effect of high-energy particles interacting with DNA ^25–27^. The present study investigated the presence of cDNMs in WGS data from the offspring of parents who had been exposed to IR either during past military service (the Radar cohort) or following the Chernobyl nuclear accident (CRU cohort of Yeager, et al., 2021). A significant increase in de novo mutation clusters was found in offspring of irradiated parents compared to controls. Furthermore, our statistical models indicated that the number cDNMs detected in offspring increased with the estimated dose of paternal IR exposure.

Most of the present statistical results indicate an influence of paternal radiation exposure on the number of cDNMs per offspring, even when only the smaller and less exposed Radar cohort is considered. Our sensitivity analysis showed that small window sizes have larger effect sizes, i.e. the difference in cDNM count between the case and control cohorts were the largest in the smaller cluster sizes (10bp - 30 bp). These observations demonstrate the lack of bias in the choice of cluster sizes and further support the hypothesis that the ROS-induced DSBs primarily result in clusters on the single- or double-digit scale. These findings are further supported by the increase of effect size and statistical significance observed in the Exposed subgroup of the Radar cohort.

In the offspring of the Radar and CRU cohort, we observed that the number of cDNMs increased by one to two per genome. To derive a clinical interpretation of these statistical results, the number and impact of cDNMs was compared with the disease burden due to all DNMs. The total number of cDNMs exceeds that of the general population by 0.6 for the Radar and 1.77 for the CRU cohort. In addition to the increase in the total number of DNMs per sample secondary to radiation exposure, it is plausible that the functional impact of cDNMs is larger compared to isolated DNMs, if they fall within coding regions of the human genome. This increased impact could lead to pathogenicity, or even embryonic lethality, in cases where cDNMs affect important parts of the coding region. However, the present authors are of the opinion that given the low overall increase in cDNMs following paternal exposure to ionizing radiation and the low proportion of the genome that is protein coding, the likelihood that a disease occurring in the offspring of exposed parents is triggered by a cDNM is minimal. Therefore, with a paternal age effect of approximately 1 additional DNM per year of paternal age, and an expected average of 60 to 80 DNMs per generation, we conclude that paternal exposure to low dose IR contributes less to an individual’s risk for genetic diseases than age. Thus, based on the current state of knowledge, the excess risk attributable to cDNMs that arose after paternal exposure to IR is negligible compared to the base risk for genetic diseases. These findings are consistent with the reports made on British nuclear test veterans, whereby no contribution to genetic disease in the offspring of former soldiers was found for potentially radiation-induced mutational patterns ^2,3^. While the expected clinical consequences of a clustered or isolated DNM is of comparable order, the consequences of a DSB that is incorrectly repaired, is usually more severe. Translocations are most likely to represent an indirect consequence of DSBs, and have been observed with increased frequency in irradiated mice as well as in the offspring of Radar soldiers ^7,25,26^. However, in contrast to cDNMs, comparing mutation rates for structural variants is more prone to errors when the respective cohorts were sequenced using different short read lengths and we have therefore refrained from assessing these statistically.

The present study had three main limitations. These concern the issues of IR dose estimation, the calling accuracy of cDNMs, and recruitment bias. Dose estimations, i.e. data on the level of exposure to IR for each soldier were retrospective and limited. In practice, that meant that radar devices that had been in active service more than 50 years ago had to be rendered operational again in order to measure scattered radiation dose profiles (Supplemental Material “Bericht S209/20”) ^32^.In addition, while the service hours and proximity to the radar device during operation and maintenance were derived from a generalized service manual of the German armies based on rank, position and mission of the soldier, in many cases the recollections of study participants differed, suggesting that this approach introduced a potential source of errors. For example, anecdotal evidence from the Radar cohort participants suggests that in contrast to official records, higher ranking soldiers participated in radar maintenance work. Thus, some of the individuals who were classified as unexposed in the present analysis, might actually have been irradiated (Figure 3 and 4), and dose values given for members of the Radar cohort are likely to have been underestimated. Additionally, since these estimations are based largely on measurements taken in a laboratory setting years after these devices have been removed from service, they should be considered inaccurate. Similar errors might be present in the CRU cohort, due to the large delay between radiation exposure and conception of most childs in their cohort, and potential inaccuracies in dose assessment ^33,55^. Despite the discussed inaccuracies of dose estimation, we proceeded with dose-effect estimations, and excluded offspring of allegedly not exposed fathers from the negative binomial regression models, including the outlier with 14 cDNMs (Figure 4). This is likely to have introduced Berksonian and classical errors into our models ^56^.

The second limitation is that a comprehensive validation of cDNM calls in all three cohorts was infeasible. We lacked DNA material to perform any validation experiments for the CRU or Inova cohorts, rendering us unable to assess the PPV of cDNM calls on this cohort independently from the Radar cohort.

A third limitation of the study was the presence of several potential ascertainment biases during recruitment. First, individuals who were under the subjective impression that they had been exposed to IR during their term of military service were more likely to participate (volunteer bias). Second, former radar soldiers who had operated devices emitting the highest quantities of stray radiation, were in their eighties at the time of recruitment. Furthermore, radar soldiers had a high personal risk for diseases following their service (survivorship bias) ^1^. Additional biases may also have arisen due to geographical effects. While all three investigated cohorts shared a common genetic ancestry, the possibility that environmental effects contributed to the observed differences, can not be ruled out ^54^. To our knowledge, the geographical origins of the three cohorts, i.e. Germany, Ukraine, and the East Coast of the USA, do not have higher than average levels of background radiation. In the present analysis, the background radiation dose was therefore assumed to be similar for all three cohorts.

The present results suggest several avenues for future research. First, studies with longer read lengths, ideally larger cohorts, and more accurate radio-dosimetry are required to improve the characterization of the dose-response relationships and disease risk of transgenerational signatures of prolonged paternal exposure to low dose IR, such as cDNMs. Second, to determine the paternal to maternal cDNM ratio in the general population, deep sequencing of an appropriate cohort using a greater read length than what was possible in the present study is required. Accurate measurement of the paternal to maternal cDNM ratio in the general population is necessary in order to enable an accurate assessment of the influence of IR exposure, since the number of clusters of paternal and maternal origin is expected to differ due to the accumulation of repair errors In exposed cohorts, a further shift towards more paternally inherited clusters would provide additional evidence for the correlation between IR exposure in the fathers and cDNM rates in the offspring. Third, modeling the gonadal dose of fathers based on the basis of cDNMs in the respective offspring could provide further interesting avenues for analysis, if the positive predictive value for cDNM detection were to improve. Currently, these models are impacted by the low sample size and the low number of true positive cDNMs per sample. However, a plausible hypothesis is that a more specific analysis would pinpoint this relationship. Fourth, subjecting samples to long read sequencing would render targeted statistical analysis of structural variants and translocations in the general population compared to exposed cohorts feasible. Fifth, further investigation of the potential impact of the linear energy transfer (LET) on the cluster size would be of interest. When individuals are subjected to IR with higher LET, the damage would be expected to increase in direct proportion to the LET level, leading to larger clusters, or an increased number of structural variants. This effect could also explain differences in the number and nature of the clusters observed between the two exposed cohorts in the present study, since the gamma ray spectrum of the IR to which the two cohorts were exposed differed, with a difference in the LET being one of the consequences thereof ^57^. Moreover, the significant difference in radiosensitivity of mature sperm and spermatogonial stem cells should be taken into account.

In conclusion, we found a significant increase in the cDNM count in offspring of irradiated parents, and a potential association between the dose estimations and the number of cDNMs in the respective offspring. Despite uncertainty concerning the precise nature and quantity of the IR involved, the present study is the first to provide evidence for the existence of a transgenerational effect of prolonged paternal exposure to low-dose IR on the human genome. The additional risk due to IR induced cDNMs on the scale of single base-pairs was very low. The present findings suggest several further promising research avenues for characterizing further transgenerational signatures of the effect of IR on the human genome, including the analysis of structural changes such as translocations, which are more complicated to detect than cDNMs.

## Data Availability

All data produced are available online at EGA with Study ID EGAS00001007321. Codes can be accessed at github (https://github.com/brand-fabian/radarstudy) and zenodo (https://doi.org/10.5281/zenodo.8431077). Supplemental Data and Supplemental Material "Bericht S209/20" is available upon request.

## Data Availability

- Sequencing data can be accessed at ega:

- EGA: Study Accession EGAS00001007321
- Code can be accessed at github and zenodo:

- https://github.com/brand-fabian/radarstudy
- https://doi.org/10.5281/zenodo.8431077

## Acknowledgements

The Radar study was funded by the German Ministry of Defense and conducted by Deutsches Zentrum für Luft und Raumfahrt (DLR) under the supervision of an expert committee consisting of Michael Krawczak, Hajo Zeeb, Ivo Gut, André Reis and experts from the Bundeswehr (Reinhard Ullmann, Matthias Port). Ionizing radiation doses for the present study were estimated by Andreas Schirmer from the “*Strahlenmessstelle der Bundeswehr”* (Radiation measurement facility of the German Federal Armed Services). The authors thank Dietmar Glaner, Josef Wiesner, and the “Bund zur Unterstützung Radargeschädigter (BzUR e.V.)” for their assistance with the recruitment of the Radar cohort. The authors also thank Hákon Jónsson from DeCode for helpful discussions on the de novo mutation rates in unexposed controls. Data was generated by the NGS core facility at Universitätsklinikum Bonn, which is part of the West German Genome Center (WGGC) and processed by the Core Unit for Bioinformatics in Bonn. Epigenetic analysis was performed by Sascha Tierling and Jörn Walter from the University Saarland. The authors thank Christine Schmäl for proofreading and language editing of the manuscript.

## Author contributions

Conceptualization: FB, HK, AK, MH, LW, DB, MN, MS, KS, PMK

Data curation: FB, AK

Formal analysis: FB, HK, LW

Funding acquisition: PMK, DB

Investigation: FB, HK, AK

Methodology: FB, HK, AK, LW, MS, PMK

Project administration: PMK

Resources: FB, AK, MH, KUL, PK, GM, DB, PMK

Supervision: DB, MN, MS, KS, PMK

Software: FB

Visualization: FB

Writing – original draft: FB, HK, AK, PMK

Writing – review & editing: FB, HK, AK, PMK

