## Supplemental Materials for "A transgenerational mutational signature from ionizing radiation exposure"

**Supplemental Material**

Fabian Brand[1], Hannah Klinkhammer[1,3], Alexej Knaus[1], Manuel Holtgrewe[2], Leonie Weinhold[3], Dieter Beule[2], Kerstin Ludwig[4], Prachi Kothiyal[5], George Maxwell[5], Markus M. Noethen[4], Matthias Schmid[3], Karl Sperling[6], and Peter M. Krawitz[1]

[1]Institut of Genomic Statistics and Bioinformatics, School of Medicine, University Bonn & University Hospital Bonn, Germany
[2]Core Unit Bioinformatics, Berlin Institute of Health, Berlin, Germany
[3]Institute of Medical Biometry, Informatics, and Epidemiology, University Hospital Bonn, Bonn, Germany
[4]Institute of Human Genetics, School of Medicine, University Bonn & University Hospital Bonn, Germany
[5]Inova Translational Medicine Institute, Inova Health System, Falls Church, VA, USA
[6]Institute of Medical and Human Genetics, Charité-Universitaetsmedizin Berlin, Germany

October 30, 2024

**A. Figures** **18**

**B. Tables** **35**

**C. Acronyms** **50**

### 1. Materials and Methods

#### 1.1. Study Cohorts

##### 1.1.1. Radar Cohort

The `Radar` cohort was recruited from former soldiers of the German Bundeswehr and East German NVA (Nationale Volksarmee). Soldiers were approached by the study team through various channels, including online networking groups, publications in reservist magazines and through the „Bund zur Unterstützung Radargeschädigter e.V.“, an organization representing soldiers which have been irradiated during their service. In total, 80 soldiers and their wives as well as 124 children could be recruited. The soldiers also supplied answers to a questionnaire which included information on their service history (i.e. the units they served with, the duration of their service and designations of radar devices in use by the unit at that time) and ID numbers of the army or armies they served with. As this is a recollection from each soldiers’ memory, we treat this information very cautiously. In addition to their service record, soldiers were also asked to disclose an abstract of their medical record, including genetic diseases and surgeries they had undergone that are potentially influenced by their army service.

For the wives and children, we collected metadata, such as their date of birth with a reduced questionnaire. Notably, this questionnaire still included their medical record in the same way as was asked from the soldiers. This could allow us to observe potential accumulations of genetic diseases in the offspring of exposed parents. All clinical data is summarized in the Supplemental Data „Clinical Data“.

All inclusion criteria were evaluated based on a questionnaire that soldiers and their offspring and wives had to fill out. Based on their answers, soldiers were only admitted to the study if there was a significant risk of exposure during their service, as judged by an independent expert. Due to the different level of detail given by different soldiers in their answers to the questionnaire, this assessment was made under the general assumption that serving in radar units of any German army until 1985 exposed soldiers to an increased dose of ionizing radiation, if they had an occupation as technician or were directly working with the radar technicians.

All participants served in one of the German armies during the period from 1950 up to the late 1990s. The West-German army recognized the threat to the health of their soldiers during the 1970s and applied technical measures to reduce the amount of stray radiation leaking from the active radar devices between 1975 and 1985.

###### Radar Cohort: Ethics Declaration

All participants of the Radarstudy were of legal age at time of recruitment and gave informed consent to the use of their data for research purposes. Ethics approval was granted by the ethics committee of the medical faculty Bonn (*Ethikkommission der Medizinischen Fakultät Bonn*). The analyses contained herein were performed within the guidelines layed out by the informed consent, within the limits of the approval granted by the ethics committee of the medical faculty Bonn for this study and adhering to all relevant laws and regulations. All identifiers were pseudonymized prior to publication and the reidentification based on identifiers mentioned in the manuscript or supplemental material is only possible by members of the research group.

##### 1.1.2. Inova Cohort

The `Inova` cohort serves as a population reference that we use to compare the genomes of the exposed cohorts against [1]. For all families of the `Inova` cohort, only the age of the parents at conception of the child is known to us. The data for this cohort was obtained in an aligned format on AWS directly from the Fairfax Medical Center. Data for the `Inova` cohort was sequenced between 2011 and 2013 on Illumina HiSeq X10 devices with a read length of 100 bp [1–3].

##### 1.1.3. Chernobyl (`CRU`) cohort

The `CRU` cohort was presented by Yeager et al. and data was obtained from dbGaP under accession `phs001163.v1.p1` [4, 5]. The original authors of the dataset report, doses in *mGy* for each father and mother. The resulting distribution of expose dose estimation values as reported by Yeager et al. is shown in Figure S1. On average, fathers were exposed to $\mu = 365.42\,\text{mGy}\,(\pm 684.55\,\text{mGy}, m = 29)$ of ionizing radiation. Mothers were less exposed at $\mu = 19.32\,\text{mGy}\,(\pm 71.64\,\text{mGy}, m = 2)$ [4, 6]. This cohort was partially sequenced on Illumina Hiseq X ($n = 214$), and Illumina NovaSeq ($n = 126$) devices.

##### 1.1.4. Parental Age

The age of the father and mother at conception of the child is an important confounder for many of the statistical analyses presented in this work. In Table S1, we show the mean paternal and maternal age for all three study cohorts, and Figure S2 shows the parental age for all offspring in this study. As expected, age of the father and age of the mother are highly correlated in all three cohorts (Pearson-R: $r = 0.71$, $p < 6 \cdot 10^{-100}$, Spearman-R: $r = 0.74$, $p < 6 \cdot 10^{-100}$).

#### 1.2. Dose Estimation

Retrospective estimation of the ionizing radiation dose each soldier was subjected to remains a challenging task. The „*Strahlenmessstelle der Bundeswehr*“ (literally: Radiation measurement facility of the Bundeswehr) analyzed the different radar devices the soldiers used and computed individual dose estimations for each soldier.

As the recollections of soldiers in our questionnaire are very inconsistent in terms of details, accuracy and length of the service description, we sent identifiers of each soldier to the Bundeswehr, after obtaining the consent from all study participants. These ID numbers allowed the Bundeswehr to find service records for all soldiers in various archives. Dose estimations were provided in a detailed report, and are based mainly on the following factors [7].

**Estimation of ionizing radiation emissions:** The local dose estimations were established for every radar device used by one of the units that a technician participating in the study served in. The amount of ionizing radiation emitted per operational hour was determined statistically based on sparse measurements dating back to the time that the respective device was in service. For some devices, especially those where experiments could be performed during operational service of the device to determine the emitted stray radiation, these estimations are expected to be accurate. Others, e.g. devices from the East-German army received little attention regarding dosimetry during their active service and the retrospective estimations therefore remain relatively inaccurate. Additional inaccuracies are expected to be introduced by the retroactive measurements taken with modified or reactivated devices, after they have long been removed from service. This is further compounded by the fact that radar systems are complex installations, where multiple sources of ionizing radiation can be found. The parts of the device emitting ionizing radiation differ based on make, model and version of any given radar device, f.e. the relevant components of the air defence system „ NIKE“ are the Magnetron of the low power acquisition radar (LOPAR), and the two Thyratrons and two Klystrons of the high power acquisition radar (HIPAR) [7].

**Estimation of working hours:** The main risk of exposure for radar technicians was during maintenance work when the radar device was active. This was necessary for example to calibrate the device (e.g. F-104 G Starfighters NASARR) or to monitor its continued operation (e.g. HAWK PAR). Based on questionnaires from earlier studies in the field, the number of hours that a technician was risking exposure given his specific military task was derived.

These two factors together can be used to obtain retrospective dose estimations. This estimation is still subject to considerable insecurities, yet it is substantially more accurate than, for example to single out specific factors from a soldiers career (e.g. Service duration, Phase of service or radar device in use at that time). We did not observe a correlation between the service duration of a given soldier and the dose estimation (Pearson-R: $r = 0.07, p = 0.43$, Spearman-R: $r = -0.12, p = 0.23$).

Using this method, Dr. Schirmer [7] reports retrospective dose estimations for 33 soldiers of the `Radar` cohort, other soldiers are unlikely to be exposed (f.e. due to their occupation, service timespan or rank). Figure S3 shows a histogram of the estimated dose. Including all zero values in the table, a radar soldier was exposed to 9.21 mSv (±53.33 mSv, $m = 0$ mSv) of stray radiation on average. We call the subset of fathers with dose $> 0$, the `EXPOSED` subcohort. In this subcohort we observe an exposure of 34.35 mSv on average (±99.77 mSv, $m = 0.0021$ mSv).

For our analysis, we exclusively used the exposure of the father together with metadata such as age at conception. To convert from *mSv* to *mGy*, we use the scaling factor of 1, under the assumption that the radar soldiers were exposed primarily to x-ray radiation emitted by relevant components of the radar devices.

#### 1.3. Primary Analysis (Sequencing)

##### 1.3.1. Whole Genome Sequencing

The `Radar` cohort was sequenced at the NGS Core Facility at the Universitätsklinikum Bonn (part of the West German Genome Center, WGGC). DNA was extracted from Blood for most samples prior to the start of the Covid-19 Pandemic. For extraction of DNA from blood, the Chemagic Magnetic Separation Module I (Chemagen, Baesweiler, Germany) was used. Later on, during the Covid-19 Pandemic, the Oragene DNA Kit (DNA Genotek Inc., Ontario, Canada) was used for processing saliva samples. To check for sample swaps and assert the correct family structure we utilized the Illumina Global Screening Array (Illumina, San Diego, USA). To construct the sequencing library, we used the TruSeq DNA PCR free Kit (Illumina, San Diego, USA) as described by the associated protocol. Afterwards, all 284 genomes were sequenced using NovaSeq 6000 sequencers to a target coverage of 30X. Sequencing was conducted in 13 batches of 25 between January 2019 and August 2020.

##### 1.3.2. PacBio Sequencing

PacBio sequencing was performed on a select subset of 55 cDNMs. Primers were designed with Primer3 for amplification of DNA fragments of three different sizes 5,000 bp, 10,000 bp or 15,000 bp containing the cDNMs and a phase informative SNP [8, 9]. Amplicon were size selected and cleaned up with AMPure XP beads utilizing SPRI technology. Amplicons were pooled, and library preparation was performed according to PacBio standard protocol for long read CCS producing HiFi reads spanning the cDNM and the associated phase informative SNP on a PacBio Sequel II.

##### 1.3.3. Sanger Sequencing

Sanger sequencing was performed on a subset of 71 cDNMs. Primers were designed with Primer3 for amplification of DNA fragments of up to 800 bp containing a cDNM and, if possible, a phase informative SNP. Amplicon size was evaluated by gel electrophoresis, cleaned and sequenced on a ABI Genetic Analyzer 3500. Data visualization was done with Geneious R9 (Biomatters).

#### 1.4. Bioinformatics Analysis

##### 1.4.1. Primary Analysis Pipeline

To provide best comparability between the study cohorts, we used the raw data from each cohort (`.fastq` for `Radar`, `.bam` for `Inova` and `CRU`), realigned it to the GRCh37 reference genome and performed a new variant calling. The data for the Inova study was provided in Glacier storage in the AWS cloud in North America. After transfer to the European data centers, the `Inova` cohort could be analyzed together with samples from the `Radar` cohort. Utilizing the Illumina DRAGEN v3.6.3 on `f1.4xlarge` Instances in the Ireland AWS datacenter, we aligned all data to the GRCh37 reference genome. This processing was orchestrated using a snakemake pipeline optimized for working with Genome data on the AWS cloud [10]. The `CRU` study data was aligned to GRCh37 and variant calling was performed using the NVIDIA Parabricks v3.7.0.1 toolkit. This computing was performed on-premises utilizing a slurm cluster featuring multiple A100 and V100S GPUs, using snakemake [11].

With either toolkit, only the alignment and Haplotypecaller jobs were performed initially. Thus, the output of the primary bioinformatics workflow consisted of alignments in `.bam` format and possible variant sites as `.gvcf` files and was downloaded from the cloud using AWS S3 Snowballs. For the final joint genotyping procedures, we used GLnexus v1.3.1 [12] with some modifications from the community to deal with DRAGEN specific VCF encodings of spanning deletions and haploid regions [12]. The GLnexus preset `gatk` was used for joint genotyping of each cohort separately. These resulting callsets were subsequently used for the *de novo* detection workflow. GLnexus was used for this step because other methods of joint genotyping (e.g. GATK CombineGVCFs and GATK JointGenotyping) struggle to scale to whole genome cohorts of the given size, and GLnexus captures a lot of the benefits of the joint calling analysis, namely much improved detection and correction of cohort or sequencer specific artifacts.

##### 1.4.2. Analysis of *de novo* Mutations

After variant calling and joint genotyping, we applied a set of filters to all three cohorts in parallel. The set of filters used is similar to the filters from Wong et al. [1]. This analysis is done using Hail v0.2.89

[13] and custom Python scripts. The usage of the Hail framework allowed us to scale the analysis and scan all three cohorts concurrently using an on-premises computing cluster. The filter criteria used are detailed in Table S2. The most impactful criterion is the allele count filter. We mandate that each *de novo* mutation does not occur in any other sample except the one it was found in. This excludes all population and familial variants that are present in any of the samples we processed. The exclusion of variants passed on inside a family is an important aspect of this filter, since the `Radar` cohort has larger families on average compared to the `CRU` and `Inova` cohorts (cf. Table S1, Figure S4). Since each *de novo* allele has to be observed exactly once in our dataset, we exclude any variant that has been passed on by the father to two or more of his children. Potentially, this can lead to scenarios where we discard true positive cDNMs clusters using this filter, thus undercounting a specific subset of potentially radiation induced mutations, but also prevents any artifacts or inherited variants to be used in the further analyses. In our analysis we also found cDNMs that failed the allele count filter $AC = 1$, where all lesions share similar population allele frequency values in common databases such as gnomAD [14]. These clusters had arisen in the preceding generation, and had been passed on by the father, but are unlikely to be radiation induced, since different mechanisms for these kinds of hotspot variants have been proposed [14]. In subsubsection 1.4.2 we report what effect this strict filtering approach has on the DNM rates that were previously reported by Yeager, *et al.* [4]. To scan the cohort VCF files for possible Mendelian errors, the Hail function `hl.de_novo` was used. This function takes the genotypes and phenotype likelihood values of the child and its parents into account to derive a posterior likelihood of the occurrence of a *de novo* event at a given location given the sequencing data of the trio.

Since the different cohorts were sequenced on multiple different Sequencers and analyzed with different calling tools, we added some more filters to correct for known sequencing artifacts. To correct for the difference between the NovaSeq and Hiseq X10 sequencers, we sequenced three families of the `Radar` cohort on both devices. This allowed us to calibrate the allele frequency filters for the differences in sequencers. We found similar mutational signatures as reported by Arora et al. in [15], in particular an increase in $C > A$ mutations. We corrected the errors by requiring $C > A$ substitutions to fulfill stricter allele frequency criteria ($0.45 < AF < 0.55$). Since the `CRU` cohort was analyzed using the GATK-equivalent Parabricks pipeline and not Illumina DRAGEN we noticed elevated ratios of $A > T$ and $G > T$ substitutions, which we corrected with an analogous allele frequency filter. With these adjustments in particular, there is a high concordance between the NovaSeq and HiSeq X callsets, for both SNVs and DNMs. The overlap of DNM calls was computed, by asserting that all DNMs in the NovaSeq data are heterozygous alternate calls in the HiSeq X10 data. If any variant failed this criterion, it was deemed False Positive. The overlap between both datasets is 90.2% in total (Table S14).

##### 1.4.3. Quality Control

We applied extensive quality control measures to ensure a consistently high quality of the sequence data. To control for sampling errors, or sample swaps in the laboratory we ran an Illumina GSA Array prior to the whole genome sequencing experiments. This allowed us to infer the cohort structure given the detected SNPs and assert that the family structure is as expected. After sequencing, we applied a standard quality control pipeline for NGS data. Our pipeline includes FastQC, mosdepth, samtools, VerifyBamID, vcftools, bcftools and peddy [16–21]. MultiQC was used to aggregate all reports from the different tools [22]. We applied samtools and mosdepth to assess the mean depth of all sequenced individuals of the `Radar` cohort and resequenced or rejected samples that had coverage substantially less than the 30X target (Cutoff: $< 28$). Using VerifyBamID, we calculated a contamination measure using data from the 1000 Genomes project as baseline. We rejected samples that had a contamination (`FREEMIX`) of more than 3%. Additionally, we checked the resulting VCF file after joint genotyping using bcftools stats for deviations from expected transition-transversion ratios. We predicted the family structure for the `Radar` cohort using vcftools relatedness2 and peddy independently. Both tools computed identical family structures for the cohorts.

Our quality control check led to the exclusion of 14 cases, that were either contaminated or had very low coverage (3X) in the initial sequencing run (Figure S5, Figure S6). All of those were sequenced from saliva samples, which were contaminated by other bacteria. No families were excluded due to the pedigree checks. However, some sampling errors were identified before sequencing and these samples were removed from all further processing and statistical analysis.

###### 1.4.4. Phasing of *de novo* Mutations

Phasing was performed for all *de novo* mutations using Unfazed and WhatsHap [23, 24]. To optimize the runtime of the tools, for each candidate *de novo* mutation, a $\pm$ 15,000 bp region was cut out from the alignments around each variant and, if necessary, realigned to the GRCh37 reference genome. The resulting VCF files from the phasing tools were distilled to a table containing the phase information for all *de novo* mutations reported in the three cohorts using custom Python scripts. This table was used in further statistical analysis and combined with the other datasets using Hail.

Factors like read length, insert size or fragment size have a big influence on the sensitivity of the phasing algorithms. Unfortunately, the `Inova` cohort has reads of length 100 bp, compared to 150 bp in both exposed cohorts (`Radar` and `CRU`). This leads to a substantial decrease in the fraction of *de novo* mutation events where the parental haplotype could be identified successfully by the chosen methods and it made comparisons between the irradiated and control cohorts challenging.

##### 1.5. Validation of clustered *de novo* mutations

Since cDNMs are known to have a higher false positive rate compared to germline variants and isolated *de novo* events, all cDNMs of the `Radar` cohort were validated by at least one of three methods. The validation process was conducted iteratively, aligned with other stages of the project to inform bioinformatics filter criteria and optimize for the detection sensitivity of cDNMs. Figure S7 details the five callsets that were established for validation purposes, and specifies the number of mutations that were validated using each of the three techniques. Broadly, each cluster was either resequenced with a different technology or analyzed in the IGV Browser to verify its existence using different algorithms and analysis tools. cDNMs were resequenced using either the Sanger or PacBio sequencing technologies (cf. subsubsection 1.3.2, subsubsection 1.3.3). Using Sanger Sequencing for validation, we analyzed a total of 71 cDNMs of which five were true positive, 39 false positive and 27 clusters failed to produce any results. During Sanger sequencing, we established and analyzed special subgroups of cDNMs that were informative for the later validation and statistical analysis efforts. We found a total of 7 different groups in the set of clusters:

- Tandem *de novo* mutations (two mutations directly next to each other)
- cDNMs with two to three lesions (within 20 bp)

- TT-GG clusters (clusters with at least two successive T>G lesions)
- Large clusters with more than 4 *de novo* mutations
- cDNMs in repetitive sequences
- cDNMs with a phase informative SNP in 250-750 bp distance
- cDNMs with a phase informative SNP in 750-15,000 bp distance

Notably, some of these groups show an increased number of false positive clusters. For example, two of 5 large clusters (two could not be evaluated) and 14 out of 15 TT-GG (one could not be evaluated) cDNMs were disproven by Sanger sequencing. The latter are sequencing artifacts, most likely due to the two-color chemistry of the Illumina NovaSeq [15]. cDNM clusters in repetitive regions of the genome also proved hard to analyze using Sanger Sequencing, since even after multiple iterations of primers, 12 of these clusters still could not be evaluated. If possible, the phase informative SNP was also analyzed with Sanger Sequencing, but due to the relatively short range (max. 800 bp), the parental origin could only be ascertained for three mutations (2 paternal, 1 maternal origin).

A further 55 cDNMs were validated using PacBio sequencing, which allows for a greater range in which the phase informative SNP for a particular cluster can be ascertained. In total, 17 cDNMs were true positive, 28 false positive and 10 could not be evaluated. Out of the 17 true positive mutations, four were found on the maternal and 9 on the paternal allele. In order to establish a positive control and to confirm the parental origin of these clusters, three clusters were validated using both Sanger and PacBio sequencing and all three clusters were confirmed by PacBio sequencing.

All cDNM clusters of the `Radar` cohort were again checked by an experienced human analyst to confirm or deny their status as true positive cluster. The following criteria are an extension of the bioinformatics filter criteria (Table S2) and were used in the analysis of cDNMs in the IGV Browser:

1. All reads spanning the lesions of a cDNM must show either all or none of the lesions of the cluster
2. Each cDNM must be covered by at least 10 reads
3. Each constituent variant of a cluster must not be found more than once in reads in the parents
4. The variant allele frequency must lie between 0.3 and 0.7
5. There must not be any sequencing artifacts within ±200 bp of the cluster
6. The cDNM must not lie within or directly (50 bp) in front of a deletion
7. The cDNM must not lie within a duplication

If one of the above-mentioned rules is broken, the variant is considered as sequencing artifact and counted as false positive. All variants that were confirmed as true positive by Sanger or PacBio sequencing also passed the inspection in the IGV Browser. Overall, after the visual assessment, 37 out of 163 clustered *de novo* mutations are found to be true positive, resulting in a positive predictive value for cDNMs of $\frac{37}{163} = 0.226$.

##### 1.5.1. Clinical analysis of validated cDNMs

After validation of all cDNMs of the Radar cohort, none of the 37 cDNMs were found in exonic coding region, 16(43%) were located in genes deeply intronic, 2(5%) were in the UTR of genes, while the majority (19, 51%) were located in intergenic regions. We classified variants in genes according to the ACMG guidelines, and found two to be likely benign as well as one variant of unknown clinical significance (Supplementary Data Validation „cDNM List“)[25–28]. None of the validated cDNMs were in association with the reported medical history of participants (cf. subsubsection 1.1.1, Supplementary Data „Clinical Data“). Although, the amount of validated cDNMs in the subsubsection 1.1.1 is very small and the mean rate of cDNMs per offspring is just 1.48 we cannot rule out that cDNMs contribute to genetic disease. Clustering of mutations may have a more significant functional impact when occurring in coding region of disease associated genes or regulatory elements than single nucleotide variants [14, 29]. This increased impact could potentially enhance their pathogenicity or potentially even lead to embryonic lethality, which would reduce their contribution to diseases that manifest later in life.

#### 1.6. Statistical Analysis

##### 1.6.1. Case-Control Matching

The parental age effect is a known confounder in our analysis. Therefore, we apply a case-control matching to create subcohorts with a homogeneous parental age distribution. To match case and control samples we use the Python package NetworkX [30]. We define three sets of nodes as $R = \{r_1, \dots, r_m\}$, $I = \{i_1, \dots i_j\}$ and $C = \{c_1, \dots c_k\}$ which represent the samples of the Radar, Inova and CRU cohorts, respectively. Let $\mathscr{X}$ be either $I$ or $C$, the set of target nodes to match against. We then define the following network for the minimum weight bipartite matching. Let the set of nodes be $N = R \cup \mathscr{X}$, and define edges $(r_m, x_k)$ for all $r_m \in R$ and $x_k \in \mathscr{X}$. Edge weights are set to

$$w(r_m, x_k) = |f(r_m) - f(x_k)| + |m(r_m) - m(x_k)|. \tag{1}$$

$m(x)$ is defined as the age of the mother of $x \in N$, $f(x)$ as the age of the father. If the age of the father or mother at conception of the child is unknown, it is imputed with the age of the partner. There are no instances in our dataset, where the age of both parents is missing. We apply the bipartite matching for $\mathscr{X} = C$ and $\mathscr{X} = I$ respectively to select a subgroup $\mathscr{X}_M \subset \mathscr{X}$ that minimizes the total age difference between the Radar cohort and one of the other cohorts at a time:

$$\mathscr{X}_M = \underset{\mathscr{X}_S \subset \mathscr{X}, |\mathscr{X}_S| = M}{\operatorname{argmin}} \sum_{r_m \in R} \sum_{x_k \in \mathscr{X}_S} w(r_m, x_k). \tag{2}$$

##### 1.6.2. Statistical Methods

We describe the characteristic traits associated with study participants using mean, median and standard deviation values or as a percentage. To assess differences in count data (e.g. number of cDNMs per sample) generalized linear models with a negative binomial distribution (negative binomial regression model (GLM)) and the log-link were applied. The model corresponds to

$$\ln(E(Y|X_1, \cdots, X_p)) = \beta_0 + \sum_{j=1}^{p} \beta_j X_j \tag{3}$$

where $Y$ denotes underlying count data and $X_1, \cdots, X_p$ are the observed variables. The estimated coefficients can be interpreted as multiplicative effects on the target. To correct for parental age as a confounder, statistical analyses were applied on age-matched data if not stated otherwise. We attempted to correct for the difference in family sizes in the three cohorts (cf. Figure S4) using a linear mixed model, but due to the small families in all cohorts these models could not estimate the variance structure and subsequently failed to converge. Under Bonferroni correction, we set a significance level of $\alpha = \frac{0.05}{9} = 0.00556$ for the statistical tests reported herein and report either nominal p-values $p_{nom}$ or adjusted p-values $p_{adj} = 9p_{nom}$.

##### 1.6.3. Analysis of *de novo* mutation rates

As detailed above, we compared the *de novo* mutation rates between the three cohorts before and after age matching. We used a negative binomial regression model with cohort affiliation as an independent variable to estimate the number of isolated *de novo* mutations (iDNMs) per sample in each of the cohorts.

Table S3 details the mean and median number of iDNMs for all three cohorts. Table S4 shows the parameters of the negative binomial regression model we used to estimate the number of iDNMs per sample. After age matching, there is no significant difference between the cohorts ($p \geq 0.4$). No change in the number of iDNMs was detected for any base exchange in the three different cohorts (Figure S8).

We also compare our data to the number of iDNMs proposed by Yeager et al. in their manuscript. Their Table 1 notes the mean number of single nucleotide *de novo* variants per sample with $72.22 \pm 13.36$. Therefore, on average we detect 6.84 iDNMs less per sample than Yeager et al., a change that can be explained by the stricter filtering criteria we apply (cf. subsubsection 1.4.2). In particular, due to the two other large case and control cohorts included in our analysis, the $AC = 1$ filter gains a substantial amount of power to exclude sequencing artifacts and common variants from the set of putative DNMs. We reckon that this is the most influential factor for the change in the mean number of iDNMs between the two analyses, followed by the changed bioinformatics pipelines and reference genome.

##### 1.6.4. Estimation of paternal age effects

To quantify and further analyze the parental age effect, we estimated the parental age effect using a negative binomial regression model. We model the number of iDNMs under the influence of the cohort, the paternal age and the interaction of these parameters. Unmatched data was used for this analysis.

Table S5 shows the estimated parameters for this model. There is no statistically significant difference between the cohorts, or the age effect in the cohorts. However, we estimate a statistically significant age effect of 2% per year of age of the father. This results in an increase of roughly 1 DNM per year, inline with other literature.

##### 1.6.5. Analysis of clustered *de novo* mutation rates

To assess the potential impact of the ionizing radiation on the children of exposed parents we analyzed the cDNM mutation rates. Table S6 shows the mean, standard deviation and median of the underlying data for all three cohorts. We modelled the number of cDNM per sample via a negative binomial regression model with cohort affiliation as an independent variable. The negative binomial regression model (Table S7) yielded the following estimates:

$$\begin{aligned}\beta_{\text{Radar}} &= 0.52 \quad p_{adj}^{\text{Radar}} = 0.045 \\ \beta_{\text{CRU}} &= 1.09 \quad p_{adj}^{\text{CRU}} < 0.001\end{aligned} \tag{4}$$

We observe significant differences among the cohorts, namely an estimated increase of 1.76 cDNM clusters on average in the `CRU` and 0.6 in the `Radar` cohort compared to the baseline `Inova` cohort ($\beta_0$).

##### 1.6.6. Simulation of positive predictive value impact

We simulated the effect of the positive predictive value for our cDNM callset using Python. Given the positive predictive value from all validation experiments, we sampled a fraction of clusters for each cohort. On the resulting dataset, we applied the negative binomial regression model to estimate the cohort effects on the number of cDNM clusters.

For the simulation we chose the parameters $ppv = 0.25$ and $n = 1000$, where $n$ is the number of simulations. Table S8 shows percentile ranges for the simulated data on age matched cohorts. The simulation experiments show stable results for p-values as well as coefficient estimates supporting the hypothesis of a significant difference in the number of cDNMs among the cohorts.

##### 1.6.7. Phasing

Phasing was performed for all *de novo* mutation events as described in subsubsection 1.4.4. The resulting data was used to assign the gamete of origin to iDNMs and cDNMs. On average, we could phase 30.35% of events in the `Radar` cohort and 31.74% of all DNMs in the `CRU` cohort, while only 6.57% could be phased in the `Inova` cohort. For both exposed cohorts, the percentage of variants that were phased to the paternal or maternal allele is very similar. Table S9 and Figure S9 detail the distribution of paternally and maternally phased *de novo* mutations for the different cohorts per sample, on non age-matched data. When considering the total number of paternally and maternally phased DNMs per cohort, we find that the numbers do not differ significantly when comparing the `CRU`(Odds-Ratio: $OR = 0.9$, Chi$^2$-Test: $p_{nom} = 0.11$) and `Radar` (Odds-Ratio: $OR = 1.11$, Chi$^2$-Test: $p_{nom} = 0.07$) cohorts to the controls. Due to the low number of phased cDNMs, in particular in the control cohort, this analysis could not be repeated for this specific set of variants.

##### 1.6.8. Subgroup Analysis

To assess the potential link to radiation for the `Radar` cohort, we performed an analysis of two different subgroups of the `Radar` cohort. We split the cohort into two groups, the `EXPOSED` (dose estimation $> 0$, $n = 30$) and `NO_EXPOSURE` (dose estimation $= 0$, $n = 77$) group. The remaining three children of two fathers could not be attributed to either group due to a lack of metadata. We compared the number of cDNM clusters for either subgroups with the control cohort. A detailed description of both subgroups can be found in Table S10. The `EXPOSED` subgroup features 1.72 cDNMs per offspring on average (Std: 1.14, Median: 1). Individuals in the `NO_EXOSURE` subgroup have 1.39 (Std: 1.85, Median: 1) cDNMs on average (cf. Figure S10). Using a negative binomial regression model the effect size is estimated at $\beta_{\texttt{NO EXPOSURE}} = 0.45$ ($p_{adj} = 0.216$) and $\beta_{\texttt{EXPOSED}}0.67$ ($p_{adj} = 0.18$) for the `NO_EXPOSURE` and `EXPOSED` subgroups respectively.

##### 1.6.9. Analysis of Cluster Sizes

Another potential effect of ionizing radiation that we can quantify is the number of lesions per cluster. It is hypothesized that the number of *de novo* mutations per cluster increases with the LET of the radiation that the father was exposed to. Clusters in the `Inova` cohort have size 2.39($\pm$0.84) (Median: 2) on average. For `Radar`, we found clusters of size 2.52($\pm$1.1) (Median: 2) and for `CRU` this value was 2.26($\pm$0.79) (Median: 2). Figure S11 shows the proportion of cDNM clusters of a given size per cohort. We modelled the cluster size dependent on the cohort affiliation using a mixed linear model with negative binomial distribution and log-link. To account for interdependencies among clusters within the same sample we incorporated a random effect with sample ID as a group variable. The resulting estimates are shown in Table S11. None of the models showed a significant change in the distribution of cluster sizes between the three cohorts in this study.

##### 1.6.10. Analysis of the radiation exposure

The base analysis in subsubsection 1.6.5 showed a large increase in the cDNM rates in both exposed cohorts. To check whether this increase in cDNMs can be explained by the dose that the parents were subjected to, we first performed another negative binomial regression model analysis, modelling the number of cDNMs given the estimated dose. In a second attempt, we estimated the same dependent variable based on the interaction of the cohort and dose estimation variables. In the both models, we purposefully excluded a dedicated parameter for the study cohort, since the `Radar` and `CRU` cohorts can be separated almost perfectly based on the dose estimations. Especially the low maximum dose in the `Radar` cohort leads to the model recognizing the cohort structure easily. This leads to the model assigning all variability to the cohort parameter, eliminating any possible insights to be gained from the dose or the interaction of both.

The first model yielded an estimation of $\beta_{\text{dose in mGy}} = 0.0005$ with $p_{adj} \leq 0.009$. The parameters of the second model are detailed in Table S12. Figure S12 shows the estimation returned by the first model against the exposed data available from both cohorts. The first model shows a significant increase in the number of cDNMs per sample with the exposure of the parents. In the second model only the `CRU` cohort shows a significant increase of cDNMs per sample dependent on the parents' exposure which is likely due to the sparse radiation estimations in the `Radar` cohort. Besides being not significant, the parameter for the `Radar` cohort is positive in the second model as well, indicating a link between the radiation exposure of the parents and the number of cDNM clusters. The estimated impact of the radiation can subsequently be calculated as

$$f(n) = \exp(\beta_0 + n \cdot \beta_1) = 1.55 \cdot e^{0.0005n},$$

where $n$ denotes the dose in $mGy$ the father was exposed to.

##### 1.6.11. Radiation exposure estimation

Subsequently, we extended this model (cf. subsubsection 1.6.10) to try to compute retrospective dose estimations based on the count of cDNMs per sample and the paternal age. As with previous models, we used the age matching method to select a subset of the control cohort to align the parental age in the case and control cohorts and fitted a gaussian regression model to estimate the potential dose of ionizing radiation a soldier was subjected to given the number of cDNMs observed. However, due to the limited amount of data and only having seven distinct input values for the cDNM count in the `CRU` cohort, the resulting model struggles to capture the variability in exposure values in both exposed cohorts (cf. Table S13a). It is therefore not suprising that the model does not show a significant connection between the cDNM count in the `CRU` cohort and the ionizing radiation exposure of the parents ($p_{adj} = 0.054$). Other configurations and excluding certain outliers (e.g. samples of the `CRU` cohort with exposure $> 1$ $Gy$) did not improve the quality of the model or its ability to predict the dose values for members of either exposed cohort. We then applied the model to the `Radar` cohort in order to gauge the potential error that was made during estimation of the radiation exposure and to validate the model performance (cf. Table S13b).

###### 1.6.12. Sensitivity analysis of cluster window sizes

In previous literature, many attempts have been made at identifying cDNMs or similar clustered mutations with different window sizes over the genome. To show the robustness of our analysis towards different sizes of clusters, we have performed the statistical analysis with four more cluster window sizes. Two of them are of similar size as the originally analyzed 20 bp, namely 10 bp and 30 bp. These window sizes have been included as a sensitivity analysis for the models of the paternal age effect (cf. subsubsection 1.6.4), the comparison of cDNM counts (cf. subsubsection 1.6.5) and the linear model estimating the number of clusters given the dose each soldier was subjected to (cf. subsubsection 1.6.10). Since these small window sizes are likely to have the same origin as 20 bp clusters, i.e. repair errors after radiation induced double strand breaks, we observe similar results as with the initial analyses. We also analyzed two larger cluster sizes, 10k bp and 47k bp, which are hypothesized to be influenced by other mechanisms, primarily maternal age effect (20k bp, [3]) and paternal age effects (46,415 bp, [31]). Figure S13 shows the model parameters, equivalent to the model in subsubsection 1.6.5. For larger cluster sizes, we observe both a drop in effect size and increase in $p_{nom}$, which is expected given that more clusters are found overall. The cDNM rates for each cohort and experiment configuration are shown in Figure S14.

[7] Andreas Schirmer. *Retrospektive Dosisberechnung Röntgenstörstrahlung*. Tech. rep. Bericht S209/20. Bundesamt für Infrastruktur, Umweltschutz und Dienstleistungen der Bundeswehr (BAIUDBw), Aug. 2021.

[30] Aric Hagberg, Pieter Swart, and Daniel S Chult. *Exploring network structure, dynamics, and function using NetworkX*. Tech. rep. Los Alamos National Lab.(LANL), Los Alamos, NM (United States), 2008.

[31] Hákon Jónsson et al. "Parental influence on human germline de novo mutations in 1,548 trios from Iceland". In: *Nature* 549.7673 (Sept. 2017), pp. 519–522. DOI: 10.1038/nature24018.

### A. Figures

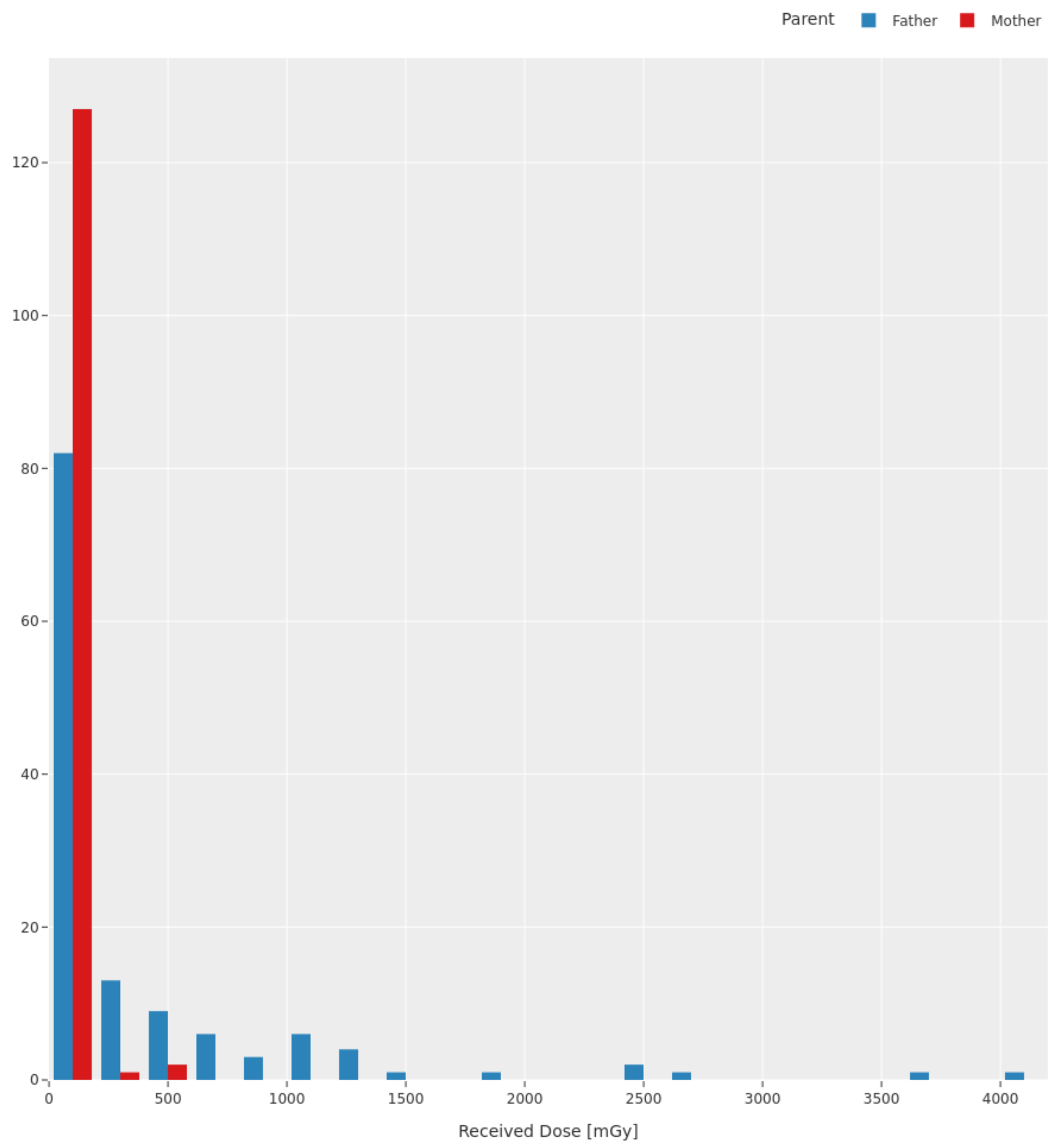

Figure S1: Exposure histogram for all families in the CRU cohort. Mothers were exposed to less radiation than fathers in general.

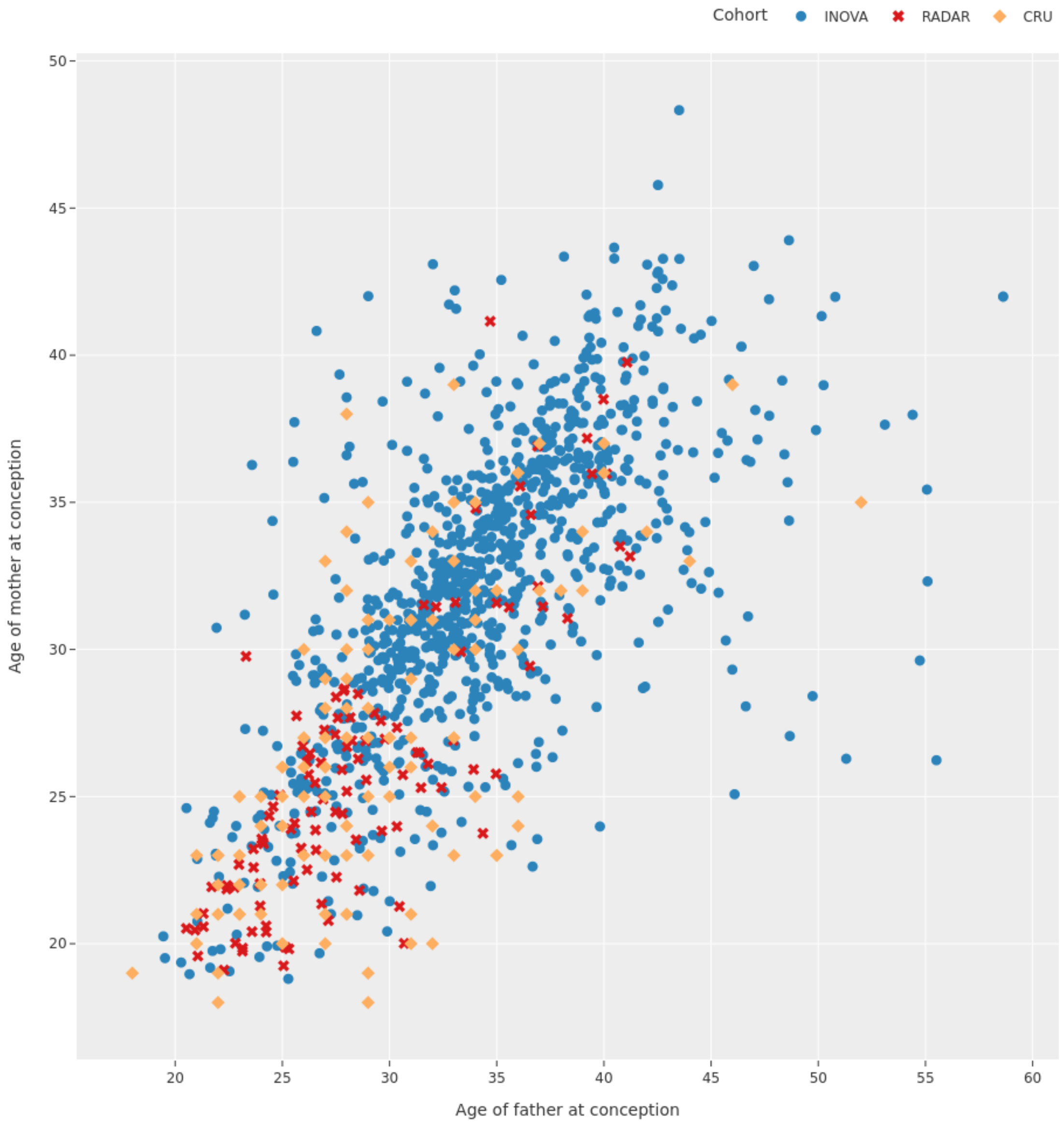

Figure S2: Age distribution of children in all three cohorts. Each point represents one child that is part of any study cohort, where the *x*-value shows the age of the father at conception of the child and the *y*-axis denotes the maternal age.

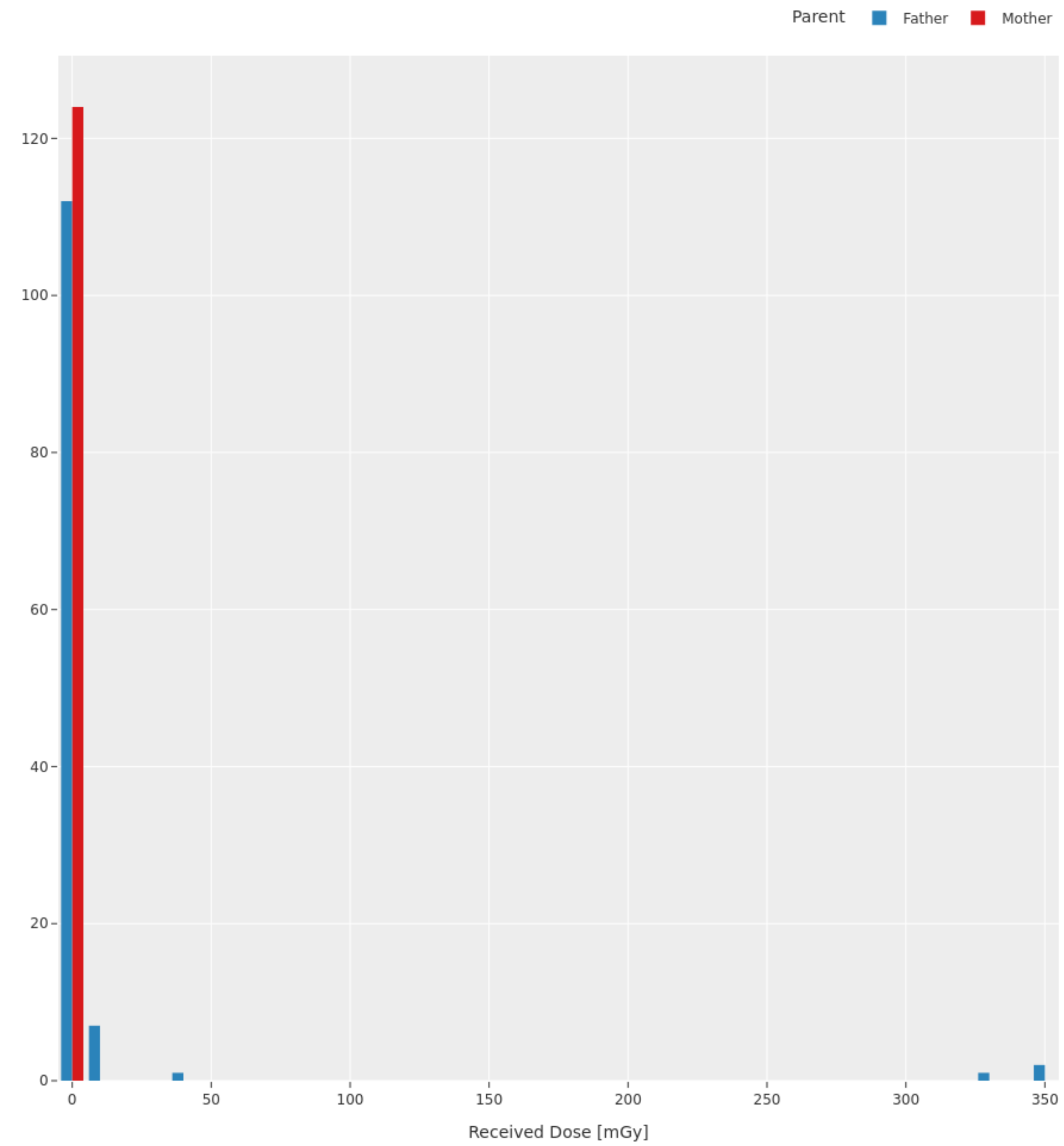

Figure S3: Exposure data from the radar cohort. Mothers are listed for completeness in the comparison to the `CRU` cohort. We could only obtain the retrospective dose estimation for 33 fathers of the radar cohort, with a maximum of 353 mGy of exposure during the service span of this particular soldier.

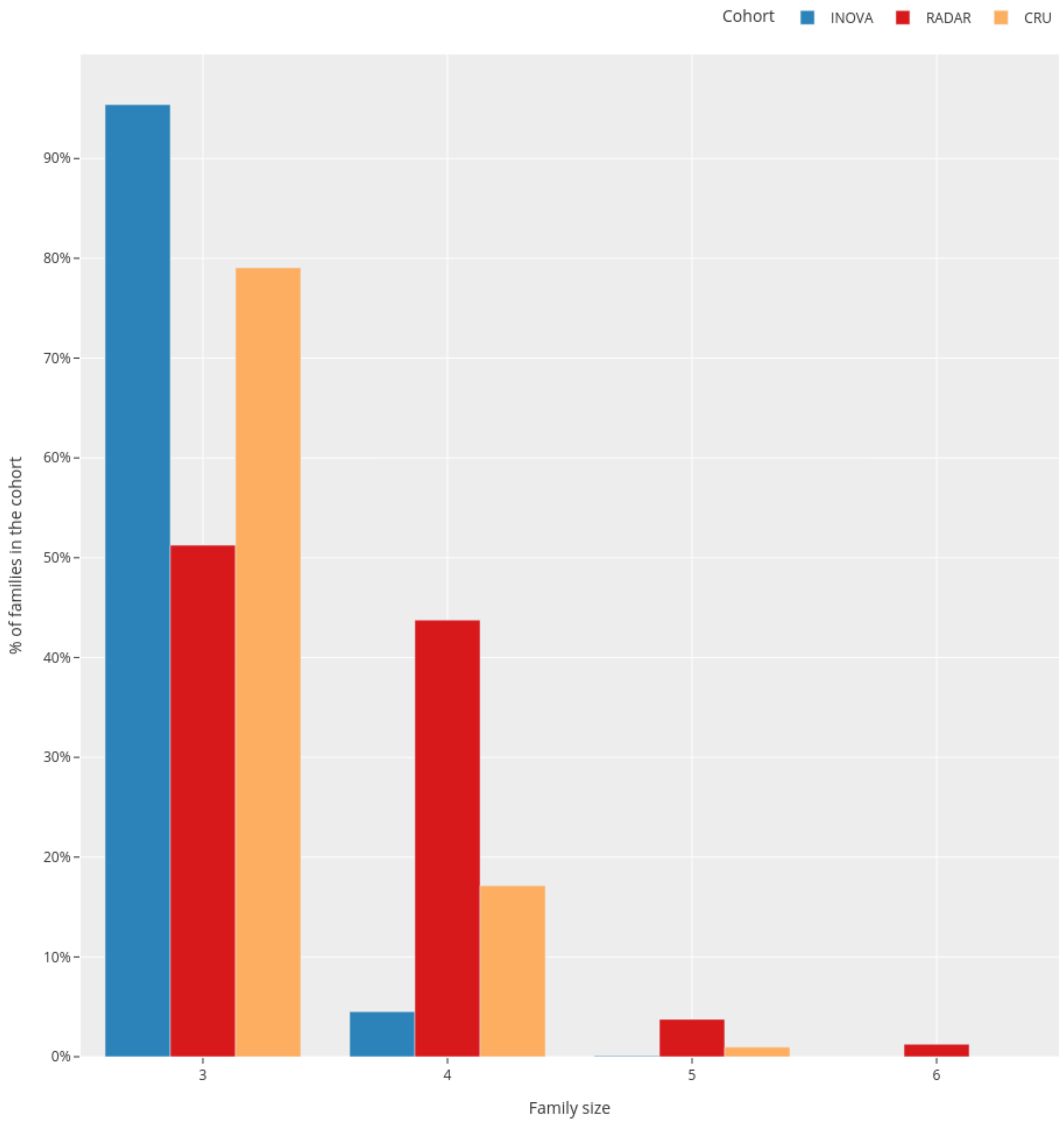

Figure S4: Bar plot of the family size distribution in each of the three analyzed cohorts. Father and Mother are included in the count for each family, so a family size of three denotes a trio family with exactly one child. The radar cohort more commonly features larger families, due to our recruitment strategy and inclusion criteria for the study.

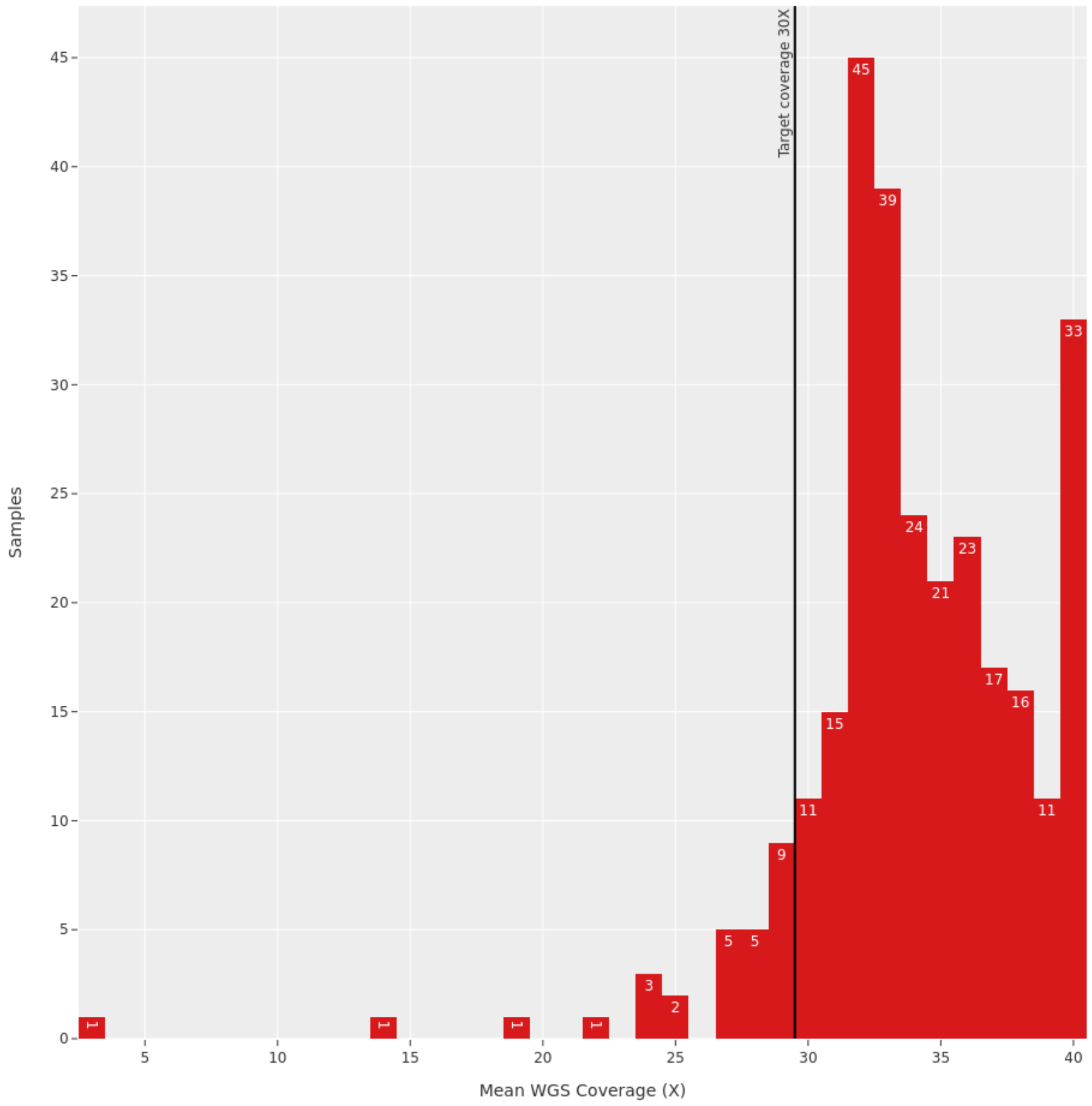

Figure S5: Coverage histogram for all samples of the `RADAR` cohort sequenced in Bonn. Samples with a whole genome coverage of $>$ 30X are considered well covered, and samples with less coverage (below a threshold of 28X) have been resequenced to meet this criterion, if at all possible.

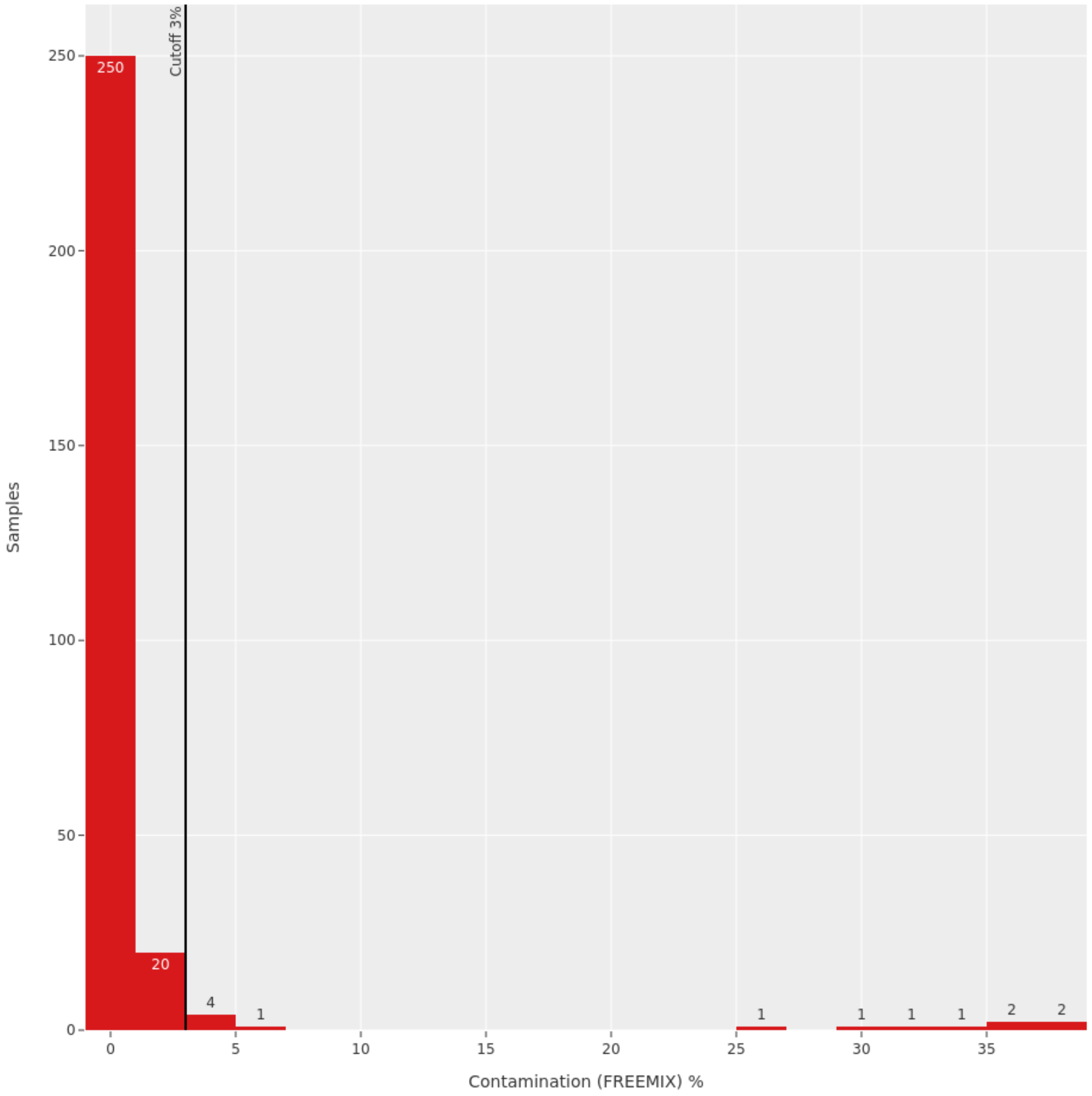

Figure S6: Contamination of `RADAR` samples with foreign DNA as computed by the tool `VerifyBamID`. A cutoff of 3% is used to identify samples which have to be resequenced or excluded from further analysis due to a high number of false positive variants introduced by foreign DNA. In total, 13 DNA samples which have been extracted from saliva fail to meet this criterion and were excluded from the following analyses.

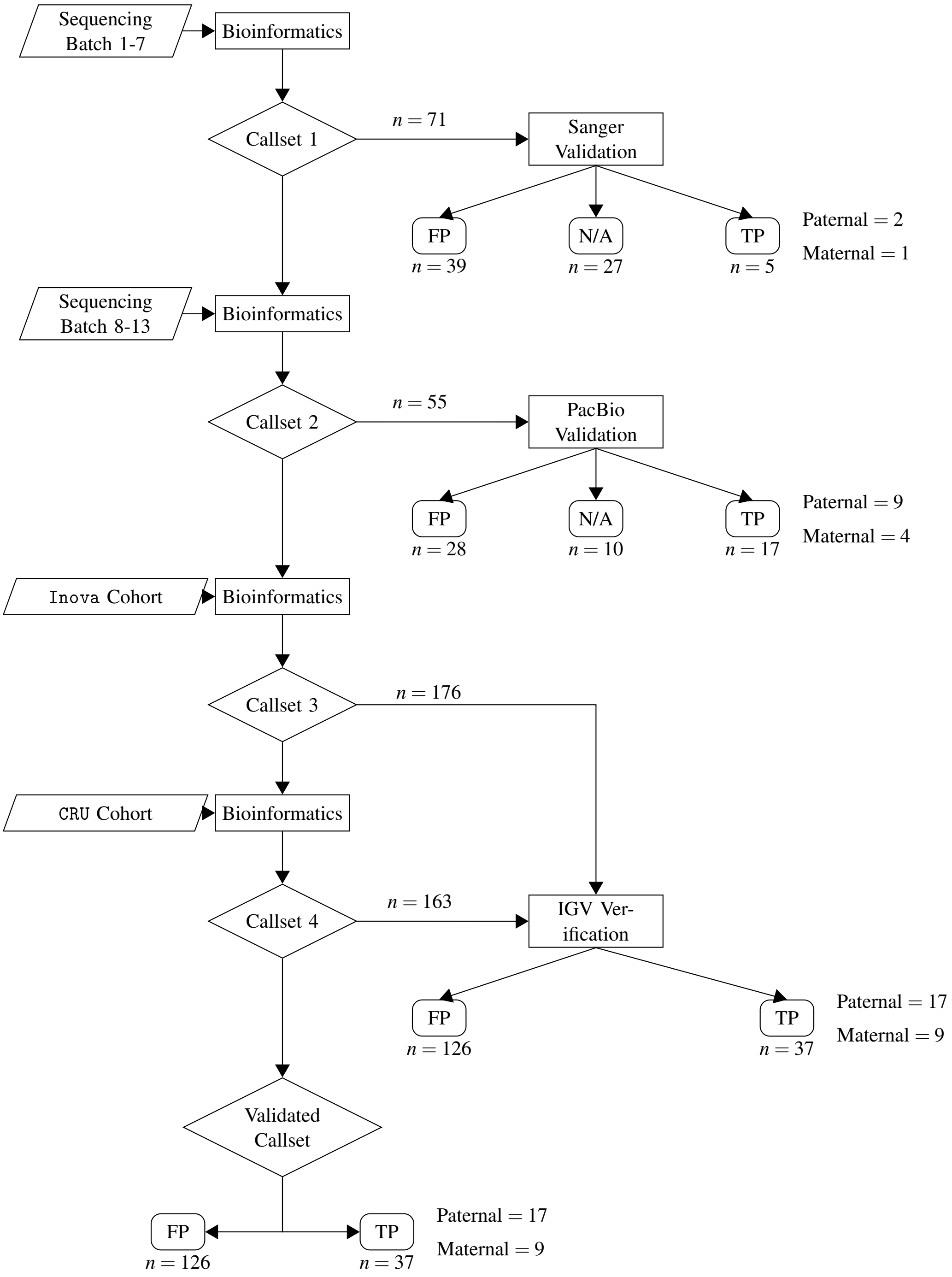

Figure S7: Visualization of the iterative process employed during the validation and verification of cD-NMs in the `Radar` cohort (cf. subsection 1.5).
The callset (diamonds) for validation changed four times during this process, after some dataset (trapezoids) was added. Each processing step (rectangle) denotes one of the processing steps that was conducted with the respective subset of one of the callsets.

Figure S7: (cont.) The result of each processing steps is denoted by the rectangles with rounded corners, which show the number of TP, N/A and FP variants for each step.

In total, 71 mutations were chosen from callset 1 for validation and resequenced using Sanger sequencing (5 TP, 39 FP, 27 N/A). 55 cDNMs from the second callset were chosen to be validated using PacBio sequencing. 12 N/A or TP variants of the 55 variants of the cDNMs selected for PacBio sequencing were also included in the previous Sanger sequencing validation. 17 out of 55 clusters were confirmed as TP cDNMs, three of which were already verified by Sanger sequencing. To build the third callset, we verified 176 cDNMs using the IGV browser. Incorporating the cDNM calls from the `Inova` (Callset 3) and `CRU` cohorts (Callset 4) always leads to a reduction in the total number of called clusters, due to variants that were earlier identified as cDNM being now reclassified and filtered based on the $AC = 1$ criterion. After inclusion of the `CRU` cohort, the number of cDNMs in callset 4 dropped to 163, the final number of cDNMs clusters. Two cDNMs that were verified in previous steps by Sanger or PacBio sequencing were now excluded due to the $AC = 1$ filter. In total, 20 more clusters could be verified using the IGV Brower. Out of the total 37 TP mutations, we could assert the parental gamete of origin in 26 cases (17 paternal, 9 maternal).

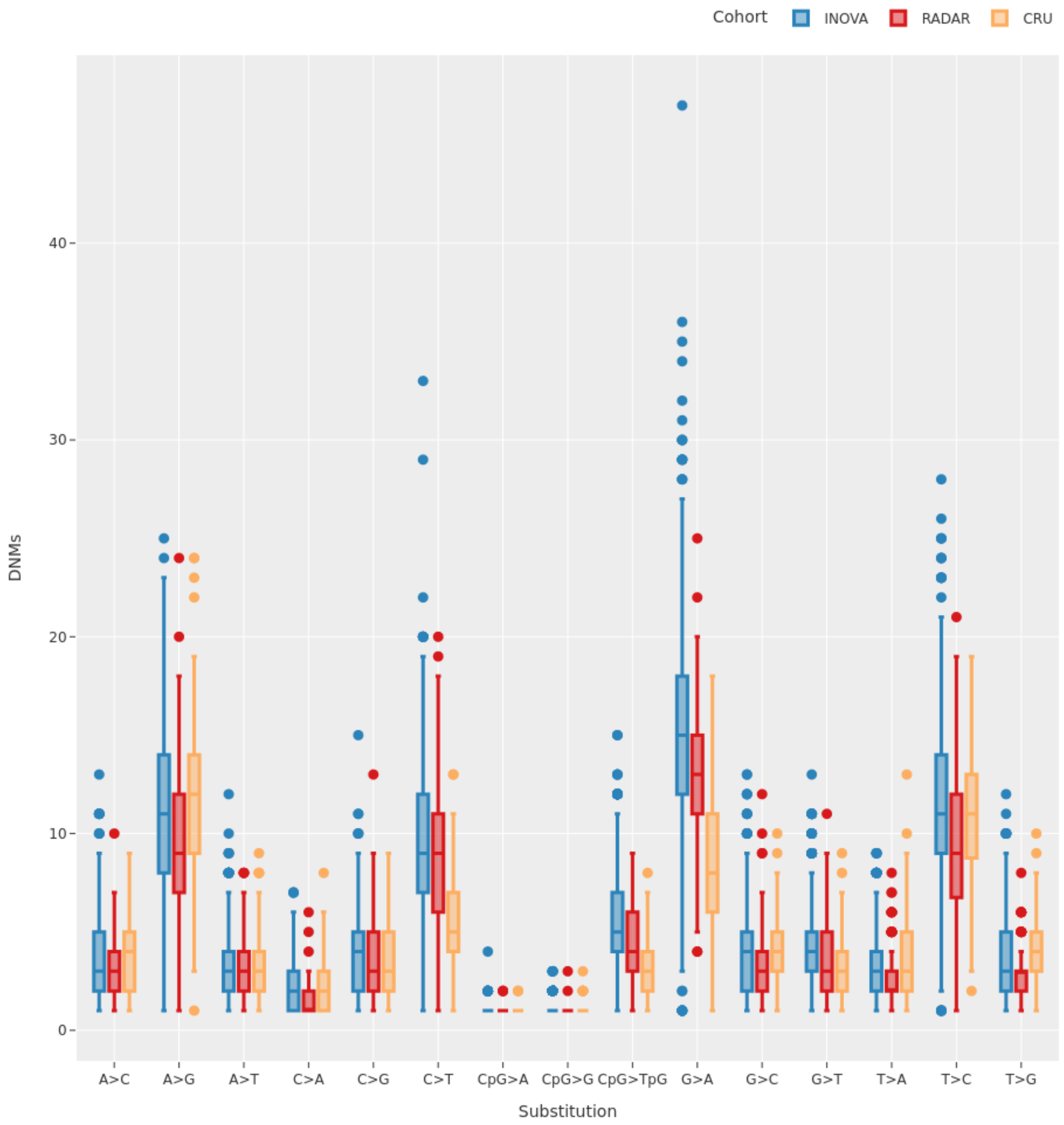

Figure S8: Substitution rates for possible base exchanges for *de novo* mutations.

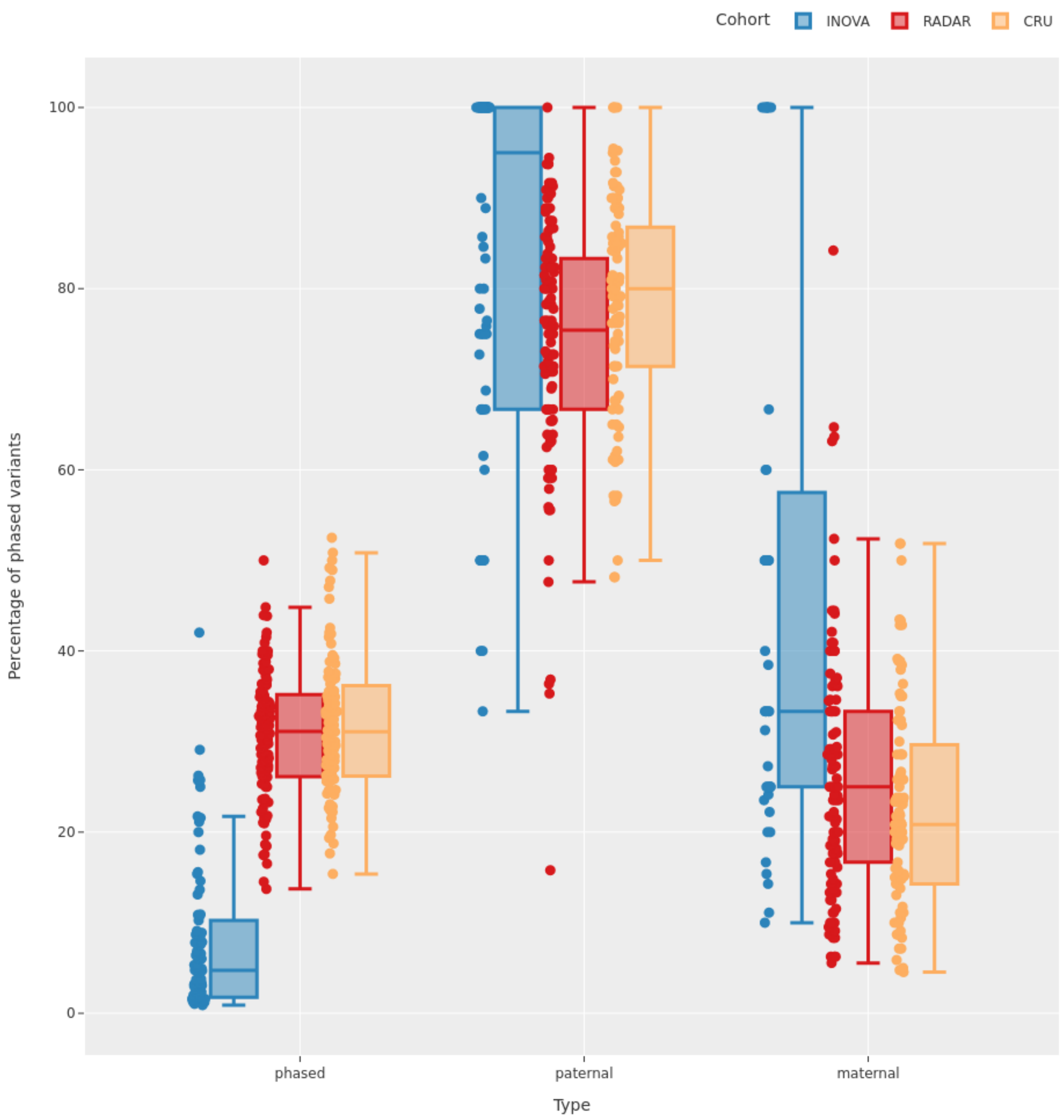

Figure S9: Percentage of *de novo* mutations phased per sample, and of those the percentage of maternally or paternally phased variants respectively. Due to the low overall percentage of phased variants in the `INOVA` cohort, many samples cluster at 100% or 0% (not shown here).

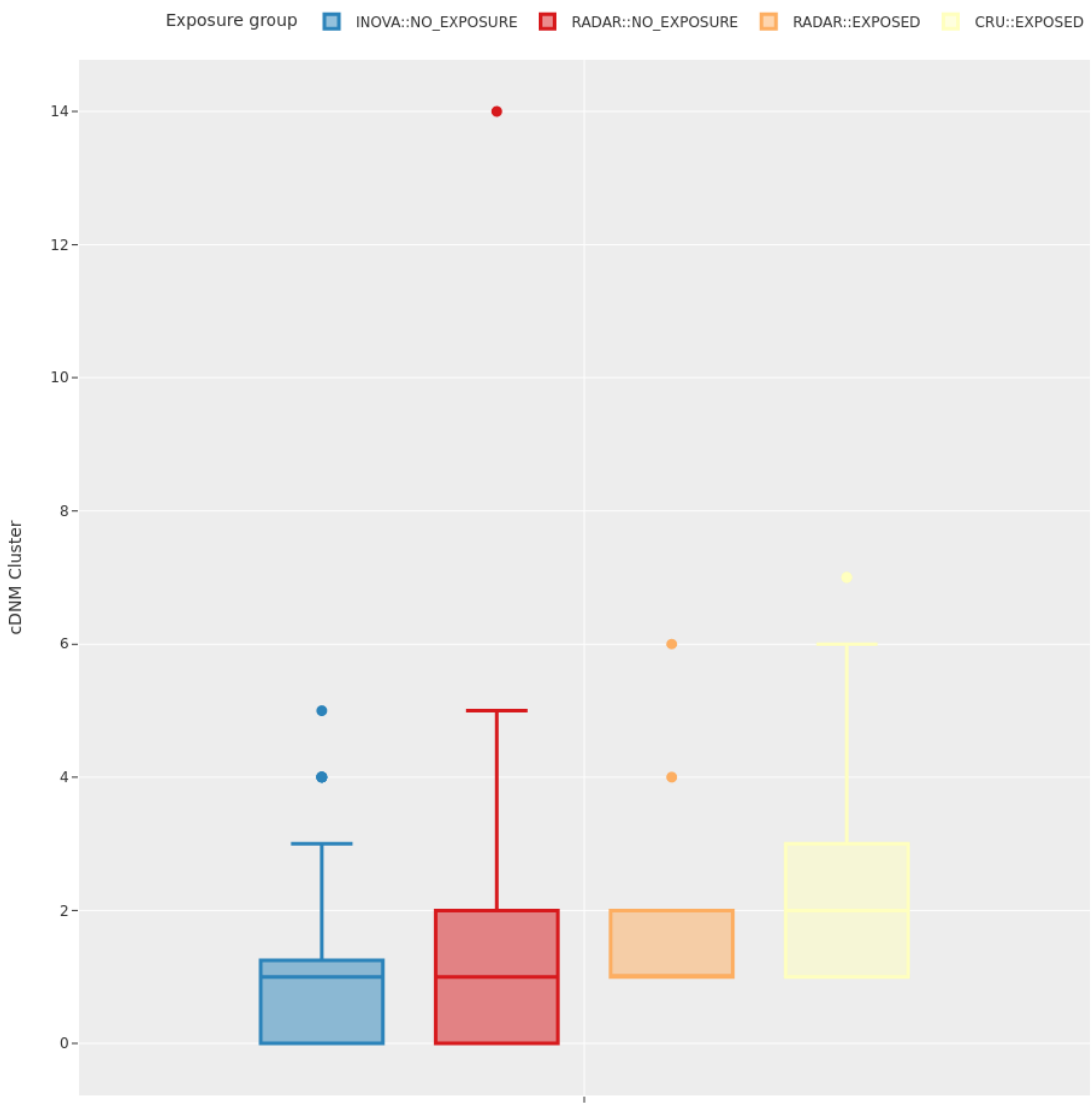

Figure S10: Number of cDNM clusters per sample grouped by the exposure status in case of the RADAR cohort. A sample is considered to be exposed to ionising radiation, if the estimated dose is greater than 0 mGy.

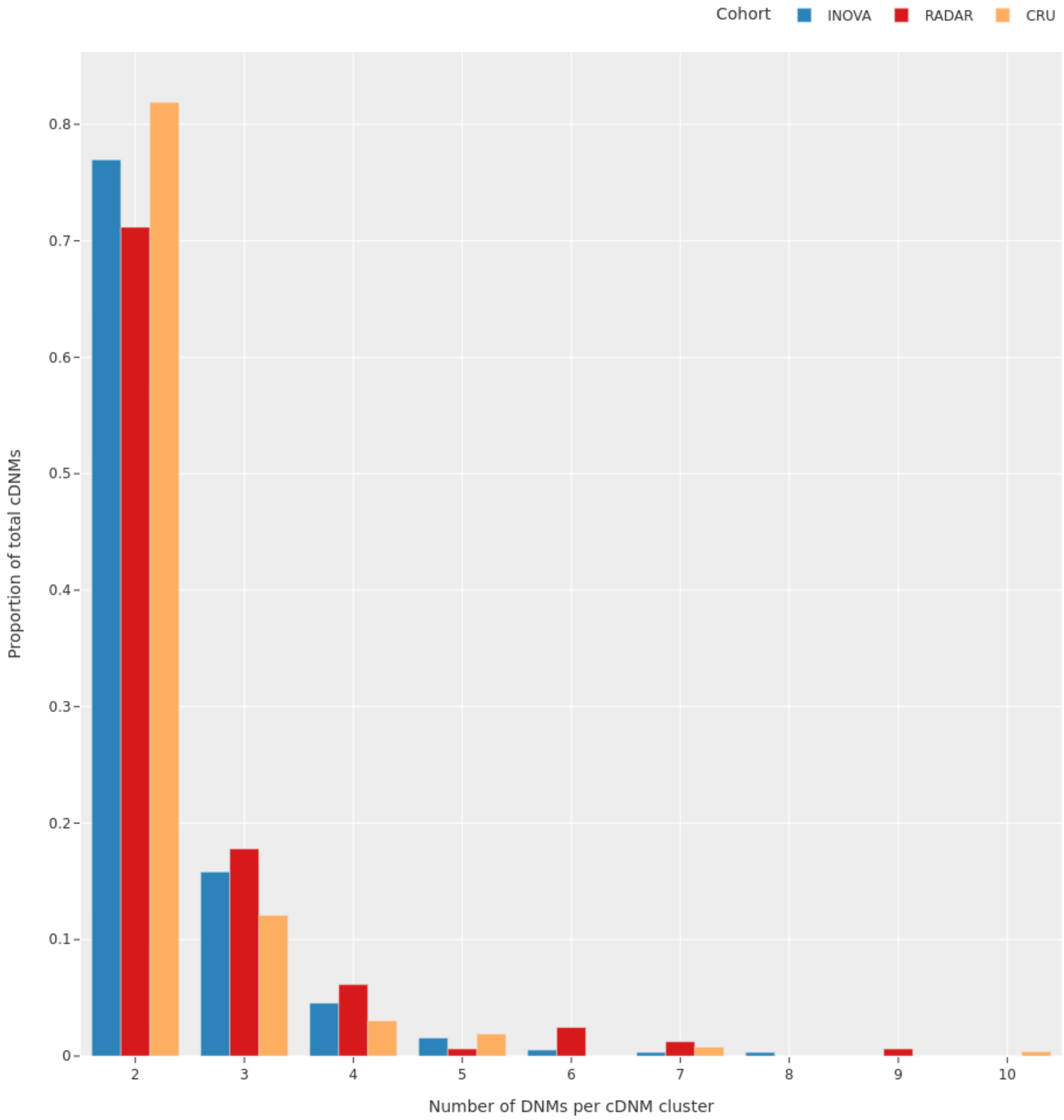

Figure S11: Size of the cDNM clusters as proportion of the total number of cDNMs observed in each cohort.

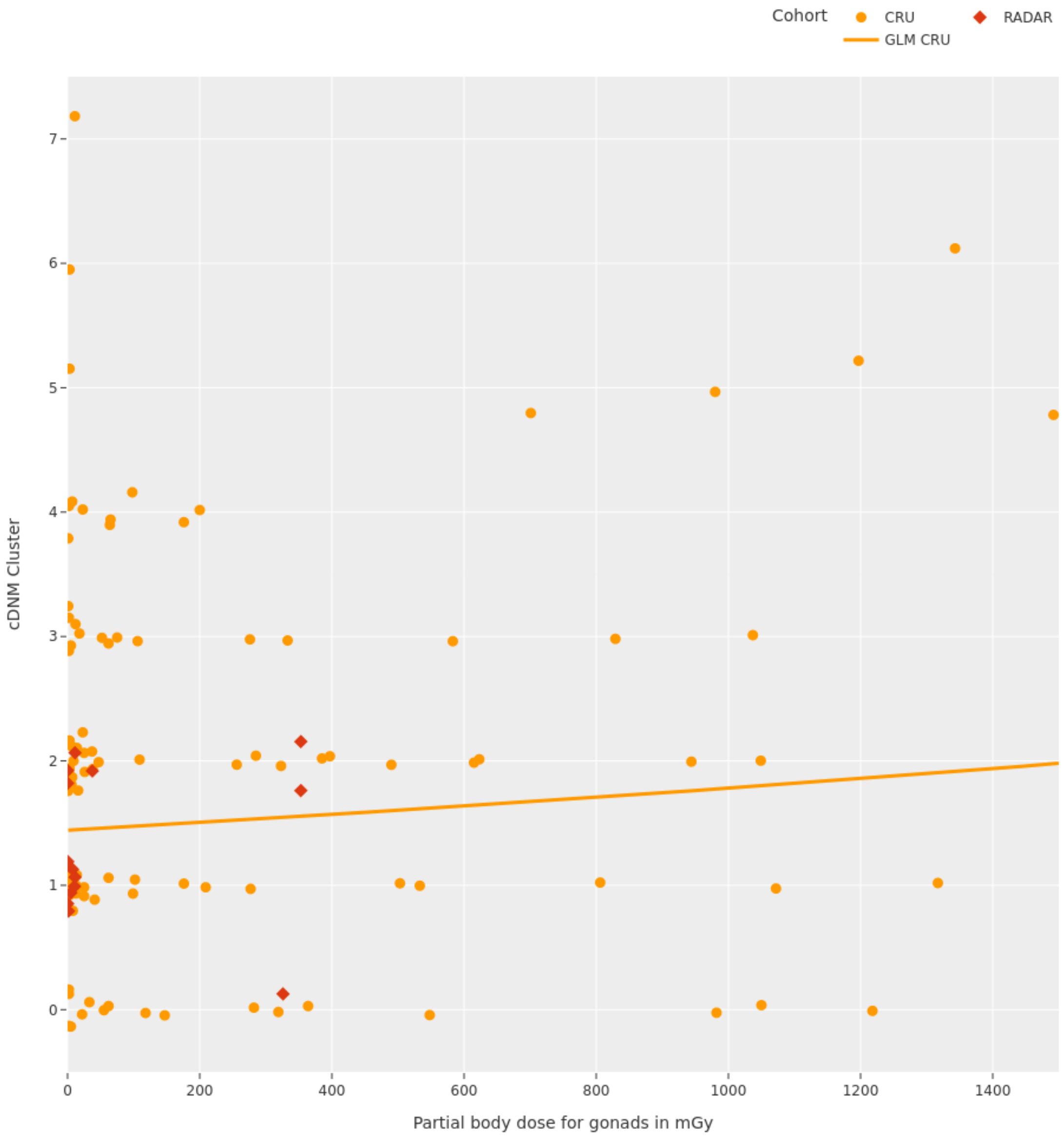

Figure S12: Estimation of the impact of the dose each soldier was subjected to on the children. The estimator is computed for each sample, given the estimated exposure of the father. No cohort specific effect was incorporated into this model, please see subsubsection 1.6.10 and Figure 4 for cohort specific effects. To be able to compute valid estimates, children from the `NO_EXPOSURE` subgroup of the radar cohort were removed from this analysis.

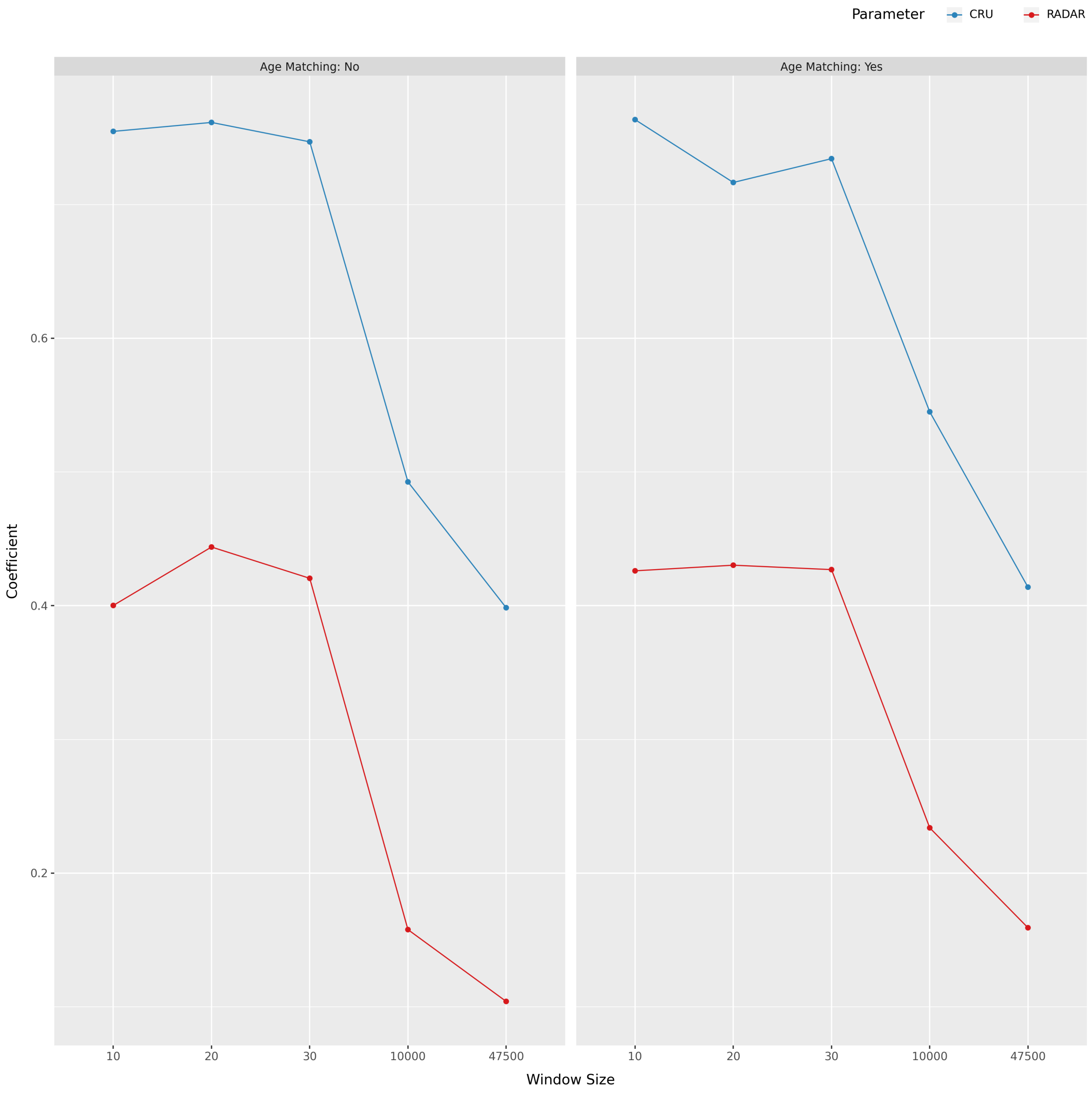

(a) Coefficients and effect sizes for the statistical model. The larger the window size, the lesser the effect we observe in both cohorts. Since more clusters are detected in general, this decline in effect size for larger window sizes is to be expected.

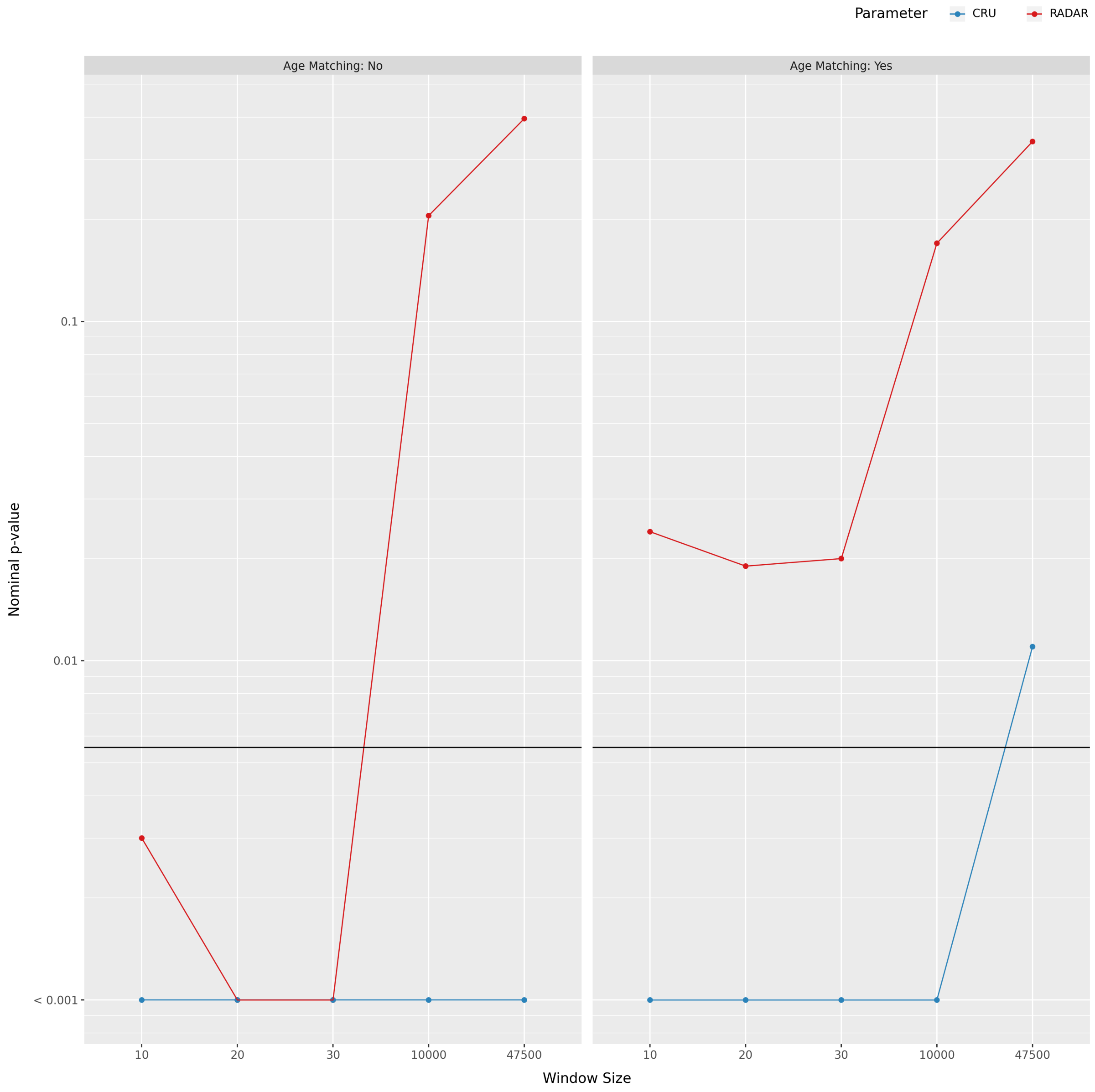

(b) Nominal $p$-values $p_{nom}$ computed by the negative binomial regression model for all five window sizes. Due to the numerical accuracy, we have cut off the y-Axis at $1 \cdot 10^{-3}$. The horizontal black bar denotes the adjusted Bonferroni corrected significance threshold $\alpha = 0.0056$

Figure S13: We conducted a sensitivity analysis with varying window sizes for cDNM detection that have previously been implicated with ionizing radiation (10, 20 and 30 bp) or maternal and general parental age effects (10k and 47.5k bp). Panel (a) shows the $\beta$ coefficients and (b) the $p_{nom}$ values for the negative binomial regression model that was built in subsubsection 1.6.5 to assess differences in cDNM rates between the study cohorts.

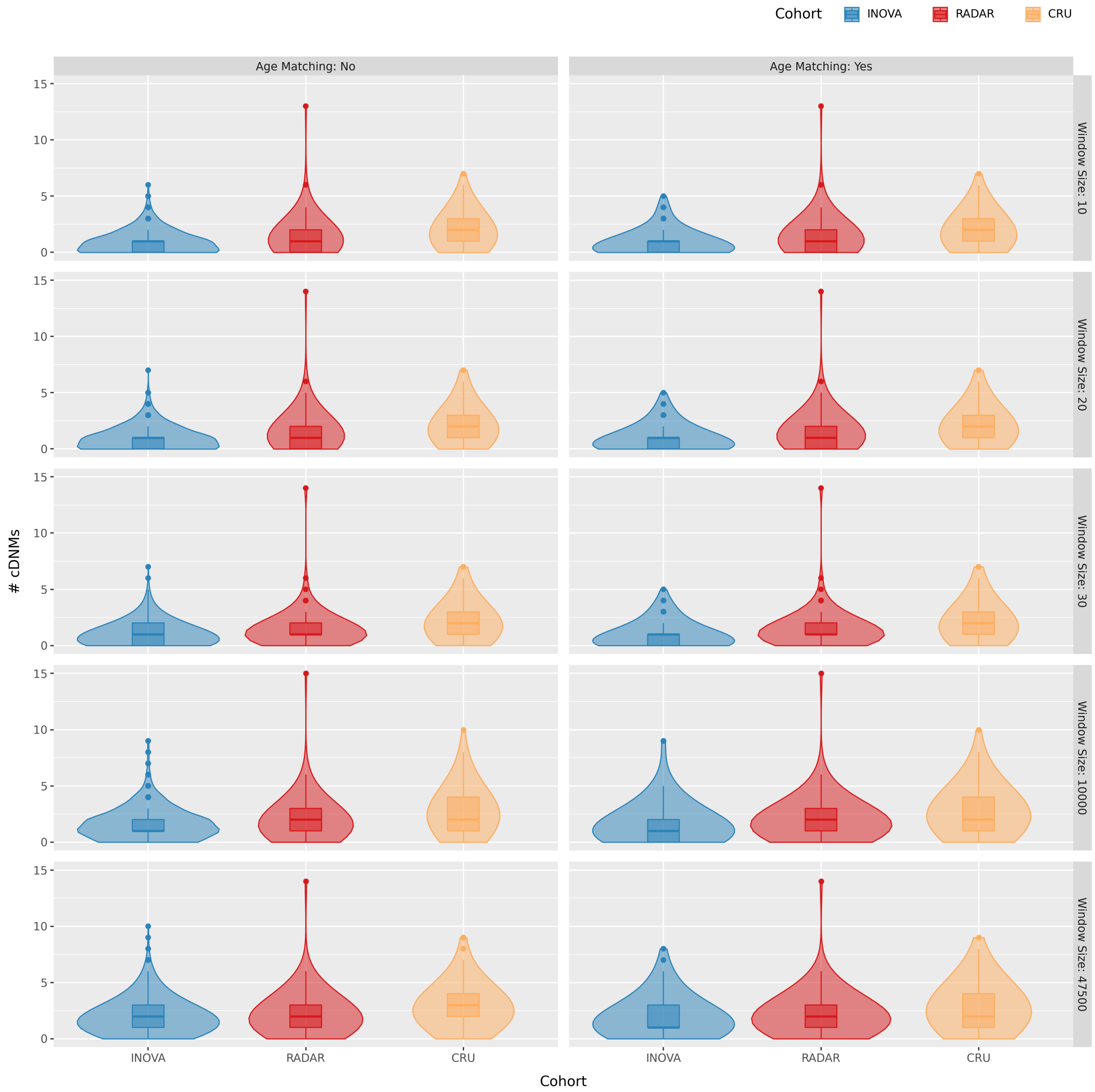

Figure S14: Violin plots for depicting the number of cDNMs per sample for each of the configurations of the sensitivity analysis. Figure 3 of the main text is equivalent to the panel „Window Size: 20“ and „Age Matching: Yes“. Inside each violin, the box plots depicts the 25%, 50% and 75% quantiles. No outliers were excluded for this analysis.

### B. Tables

| **Cohort** | # Children | # Families | Paternal Age (yrs) | Maternal Age (yrs) |
|---|---|---|---|---|
| **Radar** | 110 (46% male) | 80 | 28.54 | 25.60 |
| **CRU** | 130 (55% male) | 105 | 29.2 | 26.8 |
| **Inova** | 1275 (50% male) | 1218 | 34.50 | 32.36 |

Table S1: Cohorts analysed in the study and their respective size. The radar cohort features 1.3 children per family on average, in comparison to 1.05 (`Inova`) and 1.24 (`CRU`). The columns for paternal and maternal age show the mean age at conception in years of the fathers and mothers in the cohort. The apparent difference in sex ratios between the three cohorts, and `Radar` and `CRU` in particular is due to randomness and the small cohort sizes (cohort pairwise $\chi^2$-test: $p_{nom} \geq 0.4$).

| **Parameter** | Description | Cutoff |
|---|---|---|
| **AC** | Allele Count in all cohorts | 1 |
| **DP** | Min. Depth at variant site in index sample | 15 |
| **Parent DP** | Min. Depth at the variant site in parents | 10 |
| **AAF** | Frequency of alternate reads at the variant site | 0.3 |
| **Parent AD** | Max. number of reads with the alternate allele in parents | 1 |
| $P_{de-novo}$ | Minimum *de-novo* probability calculated by `hl.de_novo` | 0.8 |

Table S2: Filter criteria used to define the raw DNM callset. Each filter is applied to variants in child parent trio to determine the existence of such an event at a given site.

| **Group** | **Age Matched?** | # DNMs | Mean | Std. Deviation | Median | $n_{samples}$ |
|---|---|---|---|---|---|---|
| **Radar** | **No** | 7407 | 67.95 | 17.25 | 64 | 110 |
| | **Yes** | 7407 | 67.95 | 17.25 | 64 | 110 |
| **CRU** | **No** | 7192 | 65.38 | 13.76 | 65 | 129 |
| | **Yes** | 8441 | 65.43 | 13.57 | 65 | 110 |
| **Inova** | **No** | 79663 | 78.95 | 17.23 | 76 | 1009 |
| | **Yes** | 7994 | 72.67 | 18.15 | 79 | 110 |

Table S3: Number of *de novo* mutations per sample in the three study cohorts.

| | $\beta$ | $\sigma$ | z | $p_{adj}$ | [0.025 | 0.975] |
|---|---|---|---|---|---|---|
| **Intercept** | 4.2860 | 0.096 | 44.645 | 0.000 | 4.098 | 4.474 |
| **C[RADAR]** | -0.0671 | 0.136 | -0.493 | 1.000 | -0.334 | 0.200 |
| **C[CRU]** | -0.1057 | 0.136 | -0.778 | 1.000 | -0.372 | 0.160 |

Table S4: Negative binomial regression model estimating the number of *de novo* mutations per sample based on the cohort on age-matched data ($n = 330$). The intercept denotes the population background, computed based on the `Inova` cohort, **C[RADAR]** and **C[CRU]** denote effects for the `Radar` and `CRU` cohorts respectively.

| | $\beta$ | $\sigma$ | z | $p_{adj}$ | [0.025 | 0.975] |
|---|---|---|---|---|---|---|
| **Intercept** | 3.6676 | 0.192 | 19.108 | 0.000 | 3.291 | 4.044 |
| **C[RADAR]** | -0.1831 | 0.578 | -0.317 | 1.000 | -1.315 | 0.949 |
| **C[CRU]** | -0.0542 | 0.501 | -0.108 | 1.000 | -1.035 | 0.927 |
| **paternal age** | 0.0201 | 0.005 | 3.669 | 0.000 | 0.009 | 0.031 |
| **C[RADAR] : paternal age** | 0.0051 | 0.019 | 0.263 | 1.000 | -0.033 | 0.043 |
| **C[CRU] : paternal age** | -0.0009 | 0.016 | -0.057 | 1.000 | -0.033 | 0.031 |

Table S5: Negative binomial regression model estimating the paternal age effect on the number of DNMs based on 1247 observations, given the study cohort (**C[RADAR]** or **C[CRU]**) and paternal age as well as their interaction (denoted by **:**) as confounding factors.

| Group | Type | # cDNMs | Mean | Std. Deviation | Median | $n_{samples}$ |
|---|---|---|---|---|---|---|
| **Radar** | **Pairs** | 248 | 2.25 | 2.54 | 2 | 110 |
| | **Cluster** | 163 | 1.48 | 1.72 | 1 | 110 |
| **CRU** | **Pairs** | 366 | 3.33 | 3.01 | 3 | 110 |
| | **Cluster** | 291 | 2.65 | 2.19 | 2 | 110 |
| **Inova** | **Pairs** | 135 | 1.23 | 1.54 | 1 | 110 |
| | **Cluster** | 97 | 0.88 | 0.98 | 1 | 110 |

Table S6: Number of clustered *de novo* mutations per sample in the three age matched cohorts. Since a single cluster can span multiple pairs of *de novo* mutations the counts for the latter class are considerably higher.

| | $\beta$ | $\sigma$ | z | $p_{adj}$ | [0.025 | 0.975] |
|---|---|---|---|---|---|---|
| **Intercept** | -0.1258 | 0.139 | -0.903 | 1.000 | -0.399 | 0.147 |
| **C[RADAR]** | 0.5190 | 0.186 | 2.789 | 0.045 | 0.154 | 0.884 |
| **C[CRU]** | 1.0986 | 0.179 | 6.148 | 0.000 | 0.748 | 1.449 |

Table S7: Negative binomial regression model estimating the number of cDNMs per sample for all three cohorts on age matched data ($n = 330$). The control cohort Inova was used to gauge the Intercept parameters, **C[RADAR]** and **C[CRU]** report results for the respective exposed cohort.

| Group | Name | Percentile | Coefficient | $p_{nom}$ |
|---|---|---|---|---|
| **cDNM Cluster** | **RADAR** | 0.025 | 0.424883 | 0.0097 |
| | | 0.500 | 0.559616 | 0.0334 |
| | | 0.975 | 0.693147 | 0.1068 |
| | **CRU** | 0.025 | 0.904456 | $9.16 * 10^{-6}$ |
| | | 0.500 | 1.017643 | $4.34 * 10^{-5}$ |
| | | 0.975 | 1.135195 | $1.35 * 10^{-4}$ |
| | **INOVA** ($\beta_0$) | 0.025 | -1.404643 | $6.15 * 10^{-11}$ |
| | | 0.500 | -1.266493 | $4.71 * 10^{-10}$ |
| | | 0.975 | -1.174120 | $2.18 * 10^{-9}$ |

Table S8: 2.5, 50 and 97.5 Percentiles for the values retrieved from 1000 simulation runs. The coefficient `Inova` was used as Intercept $\beta_0$ in the original negative binomial regression model model, `CRU` and `Radar` are derived parameters with the associated p-value.

| **Cohort** | Allele of Origin | Mean % | Std. Deviation % | Median % |
|---|---|---|---|---|
| **Radar** | Maternal | 26.21 | 13.27 | 25 |
| | Paternal | 74.02 | 13.89 | 75.43 |
| **CRU** | Maternal | 22.34 | 11.41 | 20.42 |
| | Paternal | 78.57 | 12.02 | 80 |
| **Inova** | Maternal | 43.89 | 28.40 | 33.33 |
| | Paternal | 82.29 | 20.11 | 88.88 |

Table S9: Percentage of *de novo* mutations per sample that could be phased by the read based phasing algorithms (mean, standard deviation and median of the percentage of phased DNMs). The percentage of paternally and maternally phased DNMs is skewed in the `Inova` cohort, because of the low overall percentage of phased variants, due to the shorter read length compared to the other two cohorts. This leads to many samples that have only variants phased to one parent, shifting the distribution.

| **Cohort** | Subgroup | $n$ | Dosage Estimation | Years served |
|---|---|---|---|---|
| **Radar** | Exposed | 30 | 34.35 (±99.77) mGy | 12.69 (±10.93) y |
| | Not Exposed | 77 | N/A | 9.66 (±7.65) y |

Table S10: Characterisation of the subgroups of the `Radar` cohort used for the analysis. Due to a lack of metadata, 3 children of two fathers are excluded from this analysis. Years served denotes the average number of years served in the military in any capacity for soldiers in a given subgroup as estimated based on the answers to the questionnaire.

| | $\beta$ | $\sigma$ | z | $p_{adj}$ | [0.025 | 0.975] |
|---|---|---|---|---|---|---|
| **Intercept** | 0.8720 | 0.121 | 7.212 | 0.000 | 0.635 | 1.109 |
| **C[RADAR]** | 0.0528 | 0.152 | 0.347 | 1.000 | -0.246 | 0.351 |
| **C[CRU]** | -0.0577 | 0.140 | -0.412 | 1.000 | -0.332 | 0.217 |

Table S11: GLM estimator modelling the cluster size depending on the study cohort, for $n = 551$ clusters of varying sizes.

| | $\beta$ | $\sigma$ | z | $p_{adj}$ | [0.025 | 0.975] |
|---|---|---|---|---|---|---|
| **Intercept** | 0.4406 | 0.073 | 6.042 | 0.000 | 0.298 | 0.583 |
| **dose : C[INOVA]** | 1.1e-14 | 1.82e-15 | 6.042 | 0.000 | 7.43e-15 | 1.46e-14 |
| **dose : C[RADAR]** | 0.0007 | 0.002 | 0.291 | 1.000 | -0.004 | 0.006 |
| **dose : C[CRU]** | 0.0005 | 0.000 | 3.316 | 0.009 | 0.000 | 0.001 |

Table S12: Results of the negative binomial regression model model estimating the number of clustered *de novo* mutations based on the interaction of the estimated dose in mGy and the cohort on age matched data ($n = 330$).

| | $\beta$ | $\sigma$ | z | $p_{adj}$ | [0.025 | 0.975] |
|---|---|---|---|---|---|---|
| **Intercept** | 153.5816 | 46.770 | 3.284 | 0.009 | 61.914 | 245.249 |
| **cDNMs : C[INOVA]** | -65.6386 | 37.157 | -1.767 | 0.693 | -138.465 | 7.188 |
| **cDNMs : C[CRU]** | 61.5721 | 22.556 | 2.730 | 0.054 | 17.363 | 105.781 |

(a) Gaussian regression model fit for estimating the ionizing radiation exposure for each sample given the number of cDNMs. The model was created based on age matched data, $n = 220$.

| | | **Model** | | **Original** |
|---|---|---|---|---|
| **Cohort** | **Subcohort** | $\sqrt{\texttt{MSE}}$ $[mGy]$ | Dose Estimation $\mu, [mGy]$ | Dose Estimation $\mu, [mGy]$ |
| `CRU` | All | 672.54 | 274.31 | 365.42($\pm$684.55) |
| | $\leq 1$ $Gy$ | 277.32 | 273.84 | 160.58($\pm$248.23) |
| `Radar` | All | 264.27 | 243.31 | 9.21($\pm$53.33) |
| | `EXPOSED` | 212.36 | 235.11 | 34.35($\pm$99.77) |
| | `UNEXPOSED` | 273.93 | 244.52 | N/A |

(b) Square root of the Mean squared error of the estimations by the gaussian regression model detailed above using metadata from the `Radar` or `CRU` cohorts respectively. The Mean squared error of the `CRU` cohort is a measure of the overall accuracy of the model, since it was constructed based on this data, while results for the `Radar` cohort include other errors as well, f.e. inaccuracies in the dose reconstruction for radar soldiers. The column „Model Dose Estimation $\mu, [mGy]$“ denotes the primary output of the model, the estimated dose. For reference, the last column shows the estimates made by Yeager, *et. al* or Schirmer, *et. al*, respectively [4, 7]. Since the mean squared error of the dose estimation model is greatly affected by the outliers with large exposure, we included values for the `CRU` cohort where these 5 individuals with dose estimations exceeding 1 *Gy* were removed.

Table S13: Properties and errors for the model estimating the dose of the father, given the number of cDNMs in his children.

| Family ID | Sample ID | # DNMs | TP | FP | PPV |
|---|---|---|---|---|---|
| 1 | 1 | 57 | 49 | 8 | 0.86 |
| 2 | 2 | 72 | 65 | 7 | 0.90 |
| 2 | 3 | 68 | 67 | 1 | 0.99 |
| 3 | 4 | 63 | 52 | 11 | 0.83 |
| 3 | 5 | 66 | 63 | 3 | 0.95 |
| 3 | 6 | 54 | 48 | 6 | 0.89 |

Table S14: Positive predictive value for DNMs in six children of three families which have been sequenced on both the NovaSeq and the HiSeq. DNM calls from the NovaSeq callset have been called TP, if and only if the sample was heterozygous for this mutation in the HiSeq data. Overall, the PPV computed based on the resequenced samples is 90.2%.

### C. Acronyms

#### Glossary

**allele count** Total minor allele count observed in all samples present in the dataset. 50

**clustered *de novo* mutation** Two or more *de novo* mutations within close proximity ($\leq$ 20 bp) of each other. 50

**isolated *de novo* mutation** A single *de novo* mutation, which is not part of a cluster, i.e. there is no other *de novo* mutation within 20 bp of DNA up- or downstream. 50

**positive predictive value** Fraction of true positive observations in the total observations.. 49, 50

#### Acronyms

**AC** allele count. 50, *Glossar:* allele count

**AWS** Amazon Web Services. 4, 6

**AWS S3** Amazon Web Services Simple Storage Service. 6

**cDNM** clustered *de novo* mutation. 6–14, 18, 25, 26, 29, 30, 33–35, 41, 42, 48, 50, *Glossar:* clustered *de novo* mutation

**DNM** *de novo* Mutation. 7, 11, 12, 35, 37, 38, 40, 44, 49

**FP** False Positive. 7, 26, 49

**GLM** negative binomial regression model. 10–13, 39, 40, 42, 43, 46, 47

**iDNM** isolated *de novo* mutation. 11, 12, 50, *Glossar:* isolated *de novo* mutation

**IGV** Integrative Genomics Viewer. 8, 26

**N/A** Not Available. 26, 45, 48

**NGS** Next Generation Sequencing. 7

**PPV** positive predictive value. 49, 50, *Glossar:* positive predictive value

**SNP** Single Nucleotide Polymorphism. 6, 7, 9

**SNV** Single Nucleotide Polymorphism. 7

**TP** True Positive. 26, 49

**VCF** Variant Call Format. 6–8
